# Effectiveness and durability of mRNA-1273 BA.4/BA.5 bivalent vaccine (mRNA-1273.222) against SARS-CoV-2 BA.4/BA.5 and XBB sublineages

**DOI:** 10.1101/2023.12.11.23299663

**Authors:** Bradley K. Ackerson, Katia J. Bruxvoort, Lei Qian, Lina S. Sy, Sijia Qiu, Julia E. Tubert, Gina S. Lee, Jennifer H. Ku, Ana Florea, Yi Luo, Radha Bathala, Julie Stern, Soon K. Choi, Harpreet S. Takhar, Michael Aragones, Morgan A. Marks, Evan J. Anderson, Cindy Ke Zhou, Tianyu Sun, Carla A. Talarico, Hung Fu Tseng

## Abstract

**Background:** Emerging SARS-CoV-2 sublineages continue to cause serious COVID-19 disease, but most individuals have not received COVID-19 vaccine for >1 year. Assessment of long-term effectiveness of bivalent COVID-19 vaccines against circulating sublineages is important to inform the potential need for vaccination with updated vaccines.

**Methods:** In this test-negative study at Kaiser Permanente Southern California, sequencing-confirmed BA.4/BA.5- or XBB-related SARS-CoV-2-positive cases during 9/1/2022-6/30/2023 were matched 1:3 to SARS-CoV-2-negative controls. We assessed mRNA-1273 bivalent relative (rVE) and absolute vaccine effectiveness (VE) compared to ≥2 or 0 doses of original monovalent vaccine, respectively. Outcomes were BA.4/BA.5- or XBB-related infection, emergency department/urgent care (ED/UC) encounters, and hospitalization.

**Results:** The rVE analysis included 20,966 cases and 62,898 controls. rVE (95%CI) against BA.4/BA.5 at 14-60 days and 121-180 days was 52.7% (46.9-57.8%) and 35.5% (-2.8%-59.5%) for infection, and 59.3% (49.7-67.0%) and 33.2% (-28.2-68.0%) for ED/UC encounters. For BA.4/BA.5-related hospitalizations, rVE was 71.3% (44.9-85.1%) and 52.0% (-1.2-77.3%) at 14-60 days and 61-120 days, respectively. rVE against XBB at 14-60 days and 121-180 days, was 48.8% (33.4-60.7%) and -3.9% (-18.1-11.3%) for infection, 70.7% (52.4-82.0%) and 15.7% (-6.0-33.2%) for ED/UC encounters, and 87.9% (43.8-97.4%) and 57.1% (17.0-77.8%) for hospitalization. Results for VE and subgroup analyses (age, immunocompromise, and previous SARS-CoV-2 infection) were similar to rVE analyses.

**Conclusions:** rVE of mRNA-1273 bivalent vaccine against BA.4/BA.5 and XBB infections, ED/UC encounters, and hospitalizations waned over time. Periodic adjustment of vaccines to target emerging variants and revaccination may be important in reducing COVID-19 morbidity and mortality.

**Summary:** mRNA-1273 BA.4/BA.5 bivalent vaccine effectiveness against infection and hospitalization with BA.4/BA.5-related and XBB-related sublineages waned over time. Periodic vaccination with vaccines reflecting circulating variants may reduce SARS-CoV-2 associated morbidity and mortality.

## Introduction

As of October 7, 2023, COVID-19 has caused more than 6.4 million hospitalizations and 1.1 million deaths in the United States (U.S.) [1]. Although monovalent COVID-19 vaccines against the original SARS-CoV-2 variants were highly effective in preventing SARS-CoV-2 infections and severe outcomes, their effectiveness decreased over time due to waning immunity and emergence of immune evasive omicron variants [2]. To address this concern, updated bivalent COVID-19 vaccines containing equal amounts of the original variant and omicron BA.4/BA.5 mRNA were developed. On August 31, 2022, Moderna and Pfizer mRNA bivalent COVID-19 vaccines were authorized in the U.S. for adults aged ≥18 years who had received at least two monovalent doses [3], and by April 18, 2023, bivalent COVID-19 vaccines were authorized for all individuals aged ≥6 months [4].

Early post-authorization studies demonstrated improved effectiveness of bivalent COVID-19 vaccines against COVID-19 outcomes during BA.4/BA.5 predominance compared to original monovalent COVID-19 vaccines [5]. However, immune evasive omicron sublineages emerged, including XBB, against which several *in vitro* studies found lower neutralizing activity compared with that against previous omicron sublineages after receipt of bivalent vaccine [6]. Data on the effectiveness of bivalent COVID-19 vaccines against XBB-related sublineages indicated that receipt of bivalent vaccine provided moderately improved protection against COVID-19 compared with receipt of no COVID-19 vaccines or original monovalent vaccines only [7, 8].

However, protection waned over as little as 2-6 months, likely due to both time since vaccination and the replacement of BA.4/BA.5 by BQ.1 and XBB-related sublineages during that time. Subsequently, updated monovalent COVID-19 vaccines targeting XBB-related sublineages on September 11, 2023, for persons aged ≥6 months were authorized [9].

Despite continued high infection rates and 641,838 hospitalizations and 53,961 deaths from COVID-19 in the first 9 months of 2023, most of the US population has not received any COVID-19 vaccine in ≥1 year [1]. Since only 17% of US individuals received a bivalent vaccine dose, concerns exist that uptake of the updated monovalent XBB vaccine may also be low [10, 11]. Hence, data on the durability of bivalent COVID-19 vaccine effectiveness against COVID-19 outcomes with currently circulating variants are needed to inform regulatory agencies, healthcare providers, and individuals, of the potential importance of receiving an updated monovalent XBB vaccine [12]. Therefore, we evaluated the absolute and relative effectiveness and durability of mRNA-1273 bivalent vaccine against a range of outcomes with sequencing-confirmed omicron BA.4/BA.5- and XBB-related sublineages.

## Methods

### Study setting

Kaiser Permanente Southern California (KPSC) is a large integrated healthcare system serving over 4.8 million socio-demographically diverse members at 15 hospitals and associated medical offices across Southern California. Comprehensive electronic health records (EHRs) used for this study included demographic information, vaccinations, diagnoses, laboratory tests, procedures, and pharmacy records. External COVID-19 vaccinations were imported into members’ EHRs daily, including from the California Immunization Registry to which all COVID-19 vaccinations must be reported within 24 hours [13], and by member self-report (with valid documentation). The study was approved by the KPSC Institutional Review Board.

### Laboratory methods

Molecular diagnostic testing for SARS-CoV-2 is available to KPSC members on request for any reason and for diagnostic purposes. Specimens were primarily collected using nasopharyngeal/oropharyngeal swabs (for symptomatic or asymptomatic individuals) or saliva (for asymptomatic individuals). Specimens were tested using RT-PCR TaqPath COVID-19 High-Throughput Combo Kit (ThermoFisher Scientific). Random samples of SARS-CoV-2 positive specimens were sent weekly for whole genome sequencing (WGS), as described previously [14–16].

### Study design

We used a test-negative case-control design to assess the relative vaccine effectiveness (rVE) and absolute vaccine effectiveness (VE) of mRNA-1273 bivalent vaccine as part of a regulatory commitment from Moderna to multiple health authorities. Cases included individuals with specimens collected during 9/1/2022–6/30/2023 that tested positive by SARS-CoV-2 RT-PCR and were sent for whole genome sequencing; controls were selected from those with a negative SARS-CoV-2 RT-PCR test and no SARS-CoV-2 positive molecular or antigen test, COVID-19 diagnosis code, or antiviral treatment during the same period. Cases and controls were included if they were aged ≥6 months with ≥12 months of KPSC membership before the specimen collection (index) date or since ≥3 months of age (for ascertainment of exposure status and covariates). Individuals were excluded if they received a COVID-19 bivalent vaccine other than mRNA-1273 bivalent vaccine, received any COVID-19 vaccine <14 days before the index date or <52 days prior to the bivalent COVID-19 vaccine (≥8 weeks with a 4-day grace period), or if they had a history of COVID-19 (SARS-CoV-2 positive molecular or antigen test, COVID-19 diagnosis code, or antiviral treatment) ≤90 days before the index date. For cases or controls with more than one test meeting all criteria, only the first eligible positive or negative SARS-CoV-2 RT-PCR test was included in the analysis, respectively. Controls were randomly selected and matched 3:1 to cases by age (≤5, 6–17, 18–44, 45–64, 65–74, and ≥75 years), sex, race/ethnicity (non-Hispanic White, non-Hispanic Black, Hispanic, non-Hispanic Asian and other/unknown) and index date (±10 days). Matching was conducted separately for the rVE and VE analytic cohorts so that separate confounding adjustment and analyses could be conducted.

### Exposures

The primary exposure was receipt of mRNA-1273 bivalent vaccine (mRNA-1273.222 [original and omicron BA.4/BA.5]) following receipt of ≥2 doses of original monovalent RNA COVID-19 vaccine (Pfizer, Moderna or mixed) prior to the index date. For rVE analyses, the comparator exposure group comprised recipients of ≥2 doses of original monovalent mRNA vaccine who had not received COVID-19 bivalent vaccine prior to the index date. For VE analyses, the comparator group included individuals who had not received any COVID-19 vaccine prior to the index date.

### Outcomes

Separate analyses were conducted for rVE and VE analyses against BA.4/BA.5-related (e.g., BA.4, BA.5, BQ.1 and BQ.1.1) and against XBB-related sublineages (XBB.1.5, XBB.1.16, XBB.1.9, and others). BA.2 was not included in the analyses due to low prevalence during the study period. Outcomes assessed included SARS-CoV-2 infection (any care setting), emergency department/urgent care (ED/UC) encounter occurring on or ≤7 days after the index date, COVID-19 hospitalization, and COVID-19 hospital death. COVID-19 hospitalizations were identified among cases with index dates ≤7 days prior to or during a hospitalization with confirmation by manual chart review performed by a physician investigator (B.K.A.) and trained chart abstractors to verify the presence of severe COVID-19 symptoms.

### Covariates

Potential confounders were identified *a priori* based on the literature. Variables collected from EHRs at the index date included age, sex, self-reported race/ethnicity, socioeconomic status (Medicaid and neighborhood median household income), medical center area, and pregnancy status. Variables assessed before the index date included body mass index, smoking, Charlson comorbidity score, frailty index, chronic diseases, immunocompromised status, autoimmune conditions, and time since history of SARS-CoV-2 infection (based on available testing and diagnosis records from 3/1/2020 to the index date). To account for potential differences in care-seeking or test-seeking behaviors, health care utilization (virtual, outpatient, ED, and inpatient encounters), preventive care (other vaccinations, screenings, and wellness visits), and history of SARS-CoV-2 molecular tests were assessed. Additional covariates included month of specimen collection, specimen type, number of original monovalent COVID-19 vaccine doses prior to index date (mRNA and non-mRNA doses; for rVE analyses only) and any antiviral therapy (nirmatrelvir/ritonavir, molnupiravir, or remdesivir) ≤7 days after the index date.

### Statistical analyses

Characteristics of cases and controls for each analysis were compared using the χ^2^ test for categorical variables and Wilcoxon rank sum test for continuous variables. Absolute standardized difference (ASD) was calculated to assess the balance of covariates. Potential confounders were determined by ASD >0.1 and included in the adjusted models. Conditional logistic regression was used to estimate the adjusted odds ratios (OR) and 95% confidence intervals (CI) for vaccination with mRNA-1273 bivalent vaccine against BA.4/BA.5- and XBB-related SARS-CoV-2 infection, ED/UC encounters, COVID-19 hospitalization, and COVID-19 hospital death. We calculated rVE (%) and VE (%) as (1-OR) when OR was ≤1 and ([1/OR] – 1) when OR was >1. We also assessed rVE and VE by time since vaccination (14-60, 61-120, 121-180, and >180 days). Unconditional logistic regression was used when matching was broken. Analyses were conducted using SAS 9.4 (SAS Institute, Cary NC).

### Subgroup and sensitivity analyses

We conducted subgroup analyses by age group (≤17, 18-64, and ≥65 years), among immunocompromised individuals, and among individuals with a known history of SARS-CoV-2 infection. For sensitivity analysis, we used S-gene target failure (SGTF) status and calendar month as a proxy to assign SARS-CoV-2 lineage among cases that failed sequencing and had available SGTF data, as described previously [15, 16]. Positive specimens collected 9/1/2022–3/31/2023 with SGTF were considered to be BA.4/BA.5-related sublineages, whereas positive specimens collected 2/1/2023–6/30/2023 without SGTF were considered to be XBB-related sublineages. We also conducted rVE and VE sensitivity analyses against XBB.1.5 and against BA.4/BA.5- and XBB-related COVID-19 hospitalization and hospital death among those without antiviral treatment.

## Results

The study identified 28,227 eligible cases and 90,043 eligible controls, of which 20,966 cases and 62,898 matched controls were included in the rVE analysis, and 10,336 cases and 31,008 matched controls were included in the VE analysis (**Supplementary Figure 1**). Among 28,227 specimens sent for sequencing, 54.1% were successfully sequenced (**Supplementary Table 1, Supplementary Figures 2-3**). Compared to specimens with successful sequencing, a greater proportion of those with sequencing failure were saliva specimens (18.3% vs 8.4%) and had cycle threshold values >27 (63.2% vs 2.0%). Most variants identified were BA.4/BA.5-related (65.0%) or XBB-related sublineages (30.6%).

We describe characteristics of cases and controls included in rVE (**Table 1**) and absolute VE (**Supplementary Table 2**) analyses. In the rVE analyses, median (IQR) age in years was 52 (37-65), 57.0% were female, and 48.5% were Hispanic. In the VE analysis, median (IQR) age in years was 41 (21-61) for cases and 40 (20-61) for controls; 52.8% of both cases and controls were female, and 47.8% were Hispanic. Since most BA.4/BA.5-related cases occurred in 2022, and most XBB-related cases occurred in 2023, the average follow up time after vaccination was shorter for BA.4/BA.5-related outcomes than XBB-related outcomes (**Supplementary Figure 3)**.

**Table 1.** Characteristics of SARS-CoV-2 test-positive cases and test-negative controls with mRNA-1273 bivalent vaccine or ≥2 doses monovalent mRNA vaccines.

|  | Test Positive<br>N=20966 | Test Negative<br>N=62898 | p value | ASD |
| --- | --- | --- | --- | --- |
| Age at specimen collection date, years |  |  | 0.77 | <0.01 |
| mean (sd) | 51.58 (18.33) | 51.55 (18.63) |  |  |
| median | 52 | 52 |  |  |
| Q1, Q3 | 37, 65 | 37, 65 |  |  |
| min, max | 0.7, 102 | 0.7, 105 |  |  |
| Age at specimen collection date, years, n (%) |  |  | N/A | N/A |
| $\leq 5$ | 123 (0.6%) | 369 (0.6%) | | |
| 6-17 | 210 (1.0%) | 630 (1.0%) |  |  |
| 18-44 | 7413 (35.4%) | 22239 (35.4%) |  |  |
| 45-64 | 7857 (37.5%) | 23571 (37.5%) |  |  |
| 65-74 | 2865 (13.7%) | 8595 (13.7%) |  |  |
| $\geq 75$ | 2498 (11.9%) | 7494 (11.9%) | | |
| Sex, n (%) |  |  | N/A | N/A |
| Female | 11954 (57.0%) | 35862 (57.0%) |  |  |
| Male | 9012 (43.0%) | 27036 (43.0%) |  |  |
| Race/Ethnicity, n (%) |  |  | N/A | N/A |
| Non-Hispanic White | 4842 (23.1%) | 14526 (23.1%) |  |  |
| Non-Hispanic Black | 1763 (8.4%) | 5289 (8.4%) |  |  |
| Hispanic | 10160 (48.5%) | 30480 (48.5%) |  |  |
| Non-Hispanic Asian | 2968 (14.2%) | 8904 (14.2%) |  |  |
| Other/Unknown | 1233 (5.9%) | 3699 (5.9%) |  |  |
| Body mass index<sup>a</sup>, kg/m<sup>2</sup>, n (%) |  |  | <0.01 | 0.11 |
| <18.5 | 351 (1.7%) | 1272 (2.0%) |  |  |
| 18.5 - <25 | 4378 (20.9%) | 13618 (21.7%) |  |  |
| 25 - <30 | 6180 (29.5%) | 18983 (30.2%) |  |  |
| 30 - <35 | 4466 (21.3%) | 13887 (22.1%) |  |  |
| 35 - <40 | 2408 (11.5%) | 7015 (11.2%) |  |  |
| 40 - <45 | 1022 (4.9%) | 3130 (5.0%) |  |  |
| $\geq 45$ | 727 (3.5%) | 2211 (3.5%) | | |
| Unknown | 1434 (6.8%) | 2782 (4.4%) |  |  |
| Smoking<sup>a</sup>, n (%) |  |  | <0.01 | 0.09 |
| No | 15795 (75.3%) | 46743 (74.3%) |  |  |
| Yes | 4226 (20.2%) | 14197 (22.6%) |  |  |
| Unknown | 945 (4.5%) | 1958 (3.1%) |  |  |
| Charlson comorbidity score<sup>b,c</sup> |  |  | <0.01 | 0.10 |
| mean (sd) | 1.09 (1.94) | 1.29 (2.09) |  |  |
| median | 0 | 0 |  |  |
| Q1, Q3 | 0, 1 | 0, 2 |  |  |
| min, max | 0, 17 | 0, 17 |  |  |
| Charlson comorbidity score<sup>b,c</sup>, n (%) |  |  | <0.01 | 0.11 |
| 0 | 12596 (60.1%) | 34461 (54.8%) |  |  |
| 1 | 3537 (16.9%) | 11224 (17.8%) |  |  |
| $\geq 2$ | 4833 (23.1%) | 17213 (27.4%) | | |
| Frailty index<sup>b,d</sup> |  |  | <0.01 | 0.10 |
| mean (sd) | 0.12 (0.04) | 0.13 (0.04) |  |  |
| median | 0.11 | 0.12 |  |  |
| Q1, Q3 | 0.10, 0.14 | 0.10, 0.14 |  |  |
| min, max | 0.05, 0.40 | 0.05, 0.40 |  |  |
| Frailty index<sup>b,d</sup>, n (%) |  |  | <0.01 | 0.12 |
| Quartile 1 | 5689 (27.1%) | 15277 (24.3%) |  |  |
| Quartile 2 | 5607 (26.7%) | 15324 (24.4%) |  |  |
| Quartile 3 | 5102 (24.3%) | 15901 (25.3%) |  |  |
| Quartile 4, most frail | 4568 (21.8%) | 16396 (26.1%) |  |  |
| Chronic diseases<sup>b</sup>, n (%) |  |  |  |  |
| Kidney disease | 1796 (8.6%) | 6219 (9.9%) | <0.01 | 0.05 |
| Heart disease | 1165 (5.6%) | 4355 (6.9%) | <0.01 | 0.06 |
| Lung disease | 2602 (12.4%) | 9341 (14.9%) | <0.01 | 0.07 |
| Liver disease | 1084 (5.2%) | 3874 (6.2%) | <0.01 | 0.04 |
| Diabetes | 4093 (19.5%) | 13484 (21.4%) | <0.01 | 0.05 |
| Immunocompromised, n (%) |  |  | <0.01 | 0.09 |
| Yes | 924 (4.4%) | 4032 (6.4%) |  |  |
| HIV/AIDS | 75 | 293 |  |  |
| Leukemia, lymphoma, congenital and other immunodeficiencies, asplenia/hyposplenism | 386 | 1428 |  |  |
| Organ transplant | 127 | 483 |  |  |
| Immunosuppressant medications | 571 | 2725 |  |  |
| Autoimmune conditions<sup>b</sup>, n (%) |  |  | 0.01 | 0.02 |
| Yes | 796 (3.8%) | 2641 (4.2%) |  |  |
| Rheumatoid arthritis | 357 | 1203 |  |  |
| Inflammatory bowel disease | 148 | 569 |  |  |
| Psoriasis and psoriatic arthritis | 276 | 847 |  |  |
| Multiple sclerosis | 43 | 111 |  |  |
| Systemic lupus erythematosus | 64 | 241 |  |  |
| Pregnant at specimen collection date, n (%) |  |  | <0.01 | 0.15 |
| Yes | 331 (1.6%) | 2497 (4.0%) |  |  |
| 1st trimester | 73 | 204 |  |  |
| 2nd trimester | 100 | 262 |  |  |
| 3rd trimester | 158 | 2031 |  |  |
| History of SARS-CoV-2 infection<sup>e</sup>, n (%) |  |  | <0.01 | 0.20 |
| Yes | 5721 (27.3%) | 23002 (36.6%) |  |  |
| <180 days | 334 | 3949 |  |  |
| 180- <360 days | 1859 | 8549 |  |  |
| ≥360 days | 3528 | 10504 |  |  |
| History of SARS-CoV-2 molecular test<sup>e</sup>, n (%) | 17319 (82.6%) | 49090 (78.0%) | <0.01 | 0.11 |
| Number of outpatient and virtual visits<sup>b</sup>, n (%) |  |  | <0.01 | 0.18 |
| 0 | 830 (4.0%) | 1779 (2.8%) |  |  |
| 1-4 | 4966 (23.7%) | 11533 (18.3%) |  |  |
| 5-10 | 6027 (28.7%) | 17022 (27.1%) |  |  |
| ≥11 | 9143 (43.6%) | 32564 (51.8%) |  |  |
| Number of Emergency Department visits<sup>b</sup>, n (%) |  |  | <0.01 | 0.13 |
| 0 | 16138 (77.0%) | 44945 (71.5%) |  |  |
| 1 | 3050 (14.5%) | 11150 (17.7%) |  |  |
| ≥2 | 1778 (8.5%) | 6803 (10.8%) |  |  |
| Number of hospitalizations<sup>b</sup>, n (%) |  |  | <0.01 | 0.06 |
| 0 | 19425 (92.7%) | 57233 (91.0%) |  |  |
| 1 | 1060 (5.1%) | 3992 (6.3%) |  |  |
| ≥2 | 481 (2.3%) | 1673 (2.7%) |  |  |
| Preventive care<sup>b</sup>, n (%) | 16960 (80.9%) | 52478 (83.4%) | <0.01 | 0.07 |
| Medicaid, n (%) | 2026 (9.7%) | 7529 (12.0%) | <0.01 | 0.07 |
| Neighborhood median household income, n (%) |  |  | 0.19 | 0.02 |
| < \$40,000 | 352 (1.7%) | 1140 (1.8%) | | |
| \$40,000-\$59,999 | 3445 (16.4%) | 10066 (16.0%) | | |
| \$60,000-\$79,999 | 4974 (23.7%) | 14882 (23.7%) | | |
| ≥\$80,000 | 12148 (57.9%) | 36702 (58.4%) | | |
| Unknown | 47 (0.2%) | 108 (0.2%) |  |  |
| Medical center area<sup>f</sup> |  |  | <0.01 | 0.14 |
| Month of specimen collection, n (%) |  |  | <0.01 | 0.10 |
| September - October 2022 | 3797 (18.1%) | 12675 (20.2%) |  |  |
| November - December 2022 | 8709 (41.5%) | 23296 (37.0%) |  |  |
| January - February 2023 | 3356 (16.0%) | 11004 (17.5%) |  |  |
| March - April 2023 | 2867 (13.7%) | 9411 (15.0%) |  |  |
| May - June 2023 | 2237 (10.7%) | 6512 (10.4%) |  |  |
| Specimen type, n (%) |  |  | <0.01 | 0.10 |
| Nasopharyngeal/oropharyngeal swab | 18262 (87.1%) | 56730 (90.2%) |  |  |
| Saliva | 2704 (12.9%) | 6168 (9.8%) |  |  |
| Vaccination status, n (%) |  |  | <0.01 | 0.06 |
| Bivalent vaccinated | 3075 (14.7%) | 10712 (17.0%) |  |  |
| ≥2 doses monovalent mRNA | 17891 (85.3%) | 52186 (83.0%) |  |  |
| Number of monovalent vaccines prior to index date<sup>g</sup>, n (%) |  |  | <0.01 | 0.10 |
| 2 doses | 4283 (20.4%) | 15543 (24.7%) |  |  |
| 3 doses | 12075 (57.6%) | 33989 (54.0%) |  |  |
| ≥4 doses | 4608 (22.0%) | 13366 (21.3%) |  |  |
| Antiviral therapy within 7 days after the index date n (%) |  |  | N/A | N/A |
| Yes | 3844 (18.3%) | N/A |  |  |
| Nirmatrelvir/ritonavir | 3779 | N/A |  |  |
| Molnupiravir | 28 | N/A |  |  |
| Remdesivir | 41 | N/A |  |  |

ASD = absolute standardized difference, sd = standard deviation, N/A = not applicable
<sup>a</sup> Defined in the two years prior to specimen collection date
<sup>b</sup> Defined in the one year prior to specimen collection date
<sup>c</sup> Possible range: 0-29. Quan, H., et al. Updating and validating the Charlson comorbidity index and score for risk adjustment in hospital discharge abstracts using data from 6 countries. American Journal of Epidemiology 173.6 (2011): 676-682.
<sup>d</sup> Possible range: 0-1. Kim, D.H., et al. Measuring frailty in Medicare data: development and validation of a claims-based frailty index. The Journals of Gerontology: Series A 73.7 (2018): 980-987.
<sup>e</sup> Defined based on all available medical records from March 1, 2020 to specimen collection date
<sup>f</sup> Frequency and percent for the 19 medical center areas not shown
<sup>g</sup> Defined based on all available vaccine records from December 11, 2020 to index date

. At 14-60 days since vaccination, rVE against BA.4/BA.5 was 52.7% (46.9-57.8%) for SARS-CoV-2 infection, 59.3% (49.7-67.0%) for ED/UC encounters, and 71.3% (44.9-85.1%) for COVID-19 hospitalization (**Figure 1 and Supplementary Table 3**). However, waning of rVE against BA.4/BA.5 and XBB was observed for all outcomes >120 days. The rVE against BA.4/BA.5 was 35.5% (-2.8-59.5%) for SARS-CoV-2 infection and 33.2% (-28.2-68.0%) for ED/UC encounters at 121-180 days since vaccination. Due to insufficient numbers, rVE could not be assessed against BA.4/BA.5 for COVID-19 hospitalization at 121-180 days and for all outcomes at >180 days. rVE against XBB at 14-60 days since vaccination was 48.8% (33.4-60.7%) for SARS-CoV-2 infection, 70.7% (52.4-82.0%) for ED/UC encounters, and 87.9% (43.8-97.4%) for COVID-19 hospitalization. rVE against XBB at 121-180 days since vaccination was - 3.9% (-18.1-11.3%) for SARS-CoV-2 infection, 15.7% (-6.0-33.2%) for ED/UC encounters, and 57.1% (17.0-77.8%) for COVID-19 hospitalization. At >180 days since vaccination, point estimates of rVE against XBB were lower but had overlapping confidence intervals. Due to insufficient numbers, waning was not assessed against BA.4/BA.5 or XBB for COVID-19 hospital death; however, over the study period, confidence intervals for rVE against COVID-19 hospital death were wide and non-significant.

**Figure 1.**
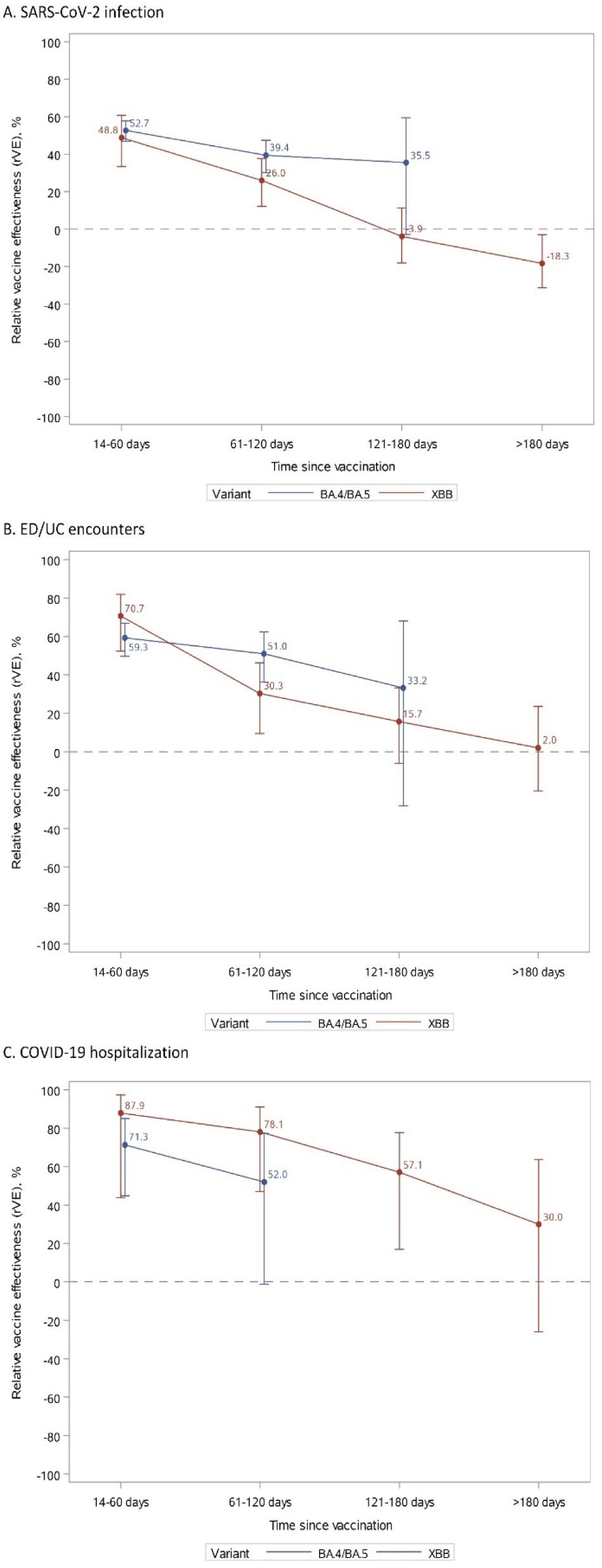
Vaccine effectiveness of mRNA-1273 bivalent vaccine vs. ≥2 monovalent mRNA vaccines against SARS-CoV-2 infection, ED/UC encounters, or COVID-19 hospitalization by variant and time since vaccination.

At 14-60 days since vaccination, VE against BA.4/BA.5 was 29.3% (19.1-38.2%) for SARS-CoV-2 infection, 55.6% (43.4-65.1%) for ED/UC encounters, and 76.9% (52.7-88.7%) for COVID-19 hospitalization (**Figure 2 and Supplementary Table 4**). A similar waning trend was observed for VE against BA.4/BA.5 and XBB against all outcomes. At 120-180 days, VE against BA.4/BA.5 was negligible for SARS-CoV-2 infection and for ED/UC encounters and could not be assessed for COVID-19 hospitalization due to insufficient numbers. VE against XBB at 14-60 days was 21.2% (-4.6-40.7%) for SARS-CoV-2 infection, 60.4% (33.8-76.3%) for ED/UC encounters, and 93.4% (68.6-98.6%) for COVID-19 hospitalization. VE against XBB quickly became negligible for SARS-CoV-2 infection and ED/UC encounters; however, at 121-180 days and >180 days since vaccination, VE against XBB for COVID-19 hospitalization was 63.8% (20.1-83.6%) and 42.4% (-19.2-73.2%), respectively.

**Figure 2.**
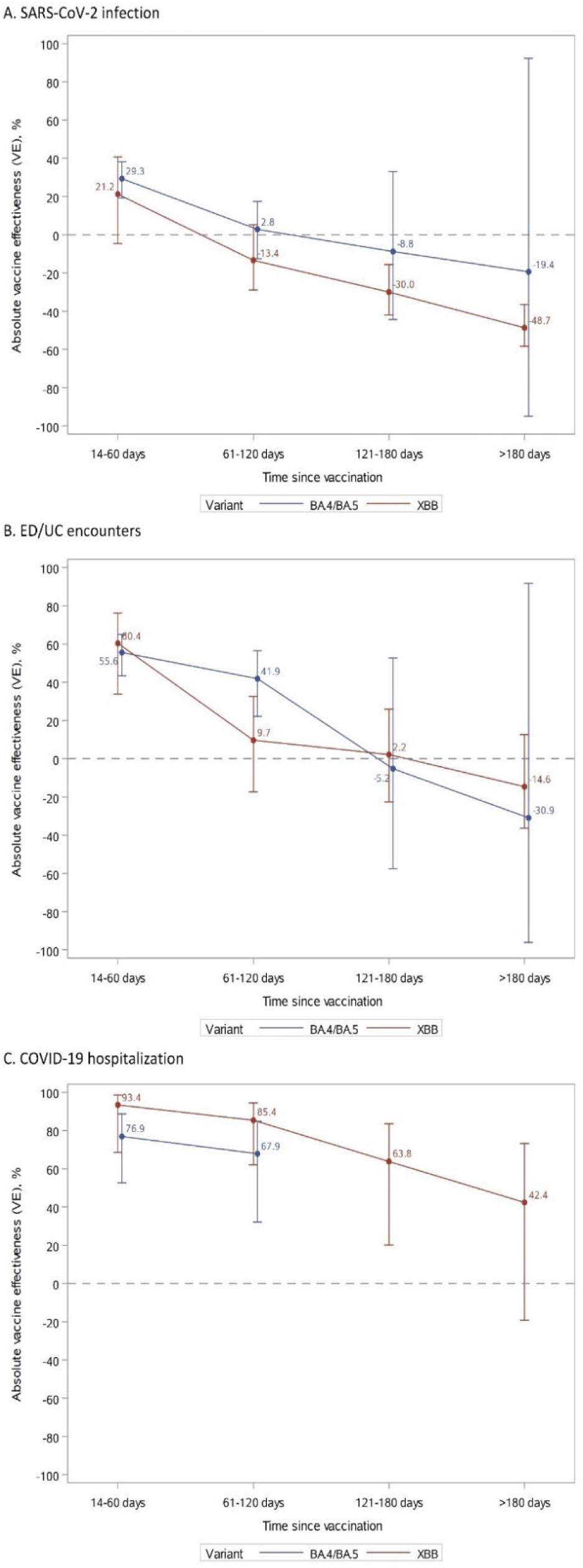
Vaccine effectiveness of mRNA-1273 bivalent vaccine vs. unvaccinated against SARS-CoV-2 infection, ED/UC encounters, or COVID-19 hospitalization by variant and time since vaccination.

Results of subgroup analyses and sensitivity analyses generally followed the main results. rVE and VE in subgroup analyses by age group, immunocompromised status, and history of SARS-CoV-2 infection were consistent the main results, although confidence intervals for subgroups analyses were wider (**Figures 3-4, Supplementary Tables 5-10)**. However, rVE against BA.4/BA.5 for ED/UC encounters was lower among immunocompromised individuals (12.1% [-33.1-48.4%]), and VE against BA.4/BA.5 for SARS-CoV-2 infection was higher among individuals with previous SARS-CoV-2 infection (51.8% [37.5-62.8%]). Results were similar for sensitivity analyses using SGTF data to assign unidentified subvariants, sensitivity analyses against XBB.1.5 specifically, and sensitivity analyses against BA.4/BA.5 and XBB for COVID-19 hospitalization and hospital death among those without antiviral treatment (**Supplementary Tables 11-16**).

**Figure 3.**
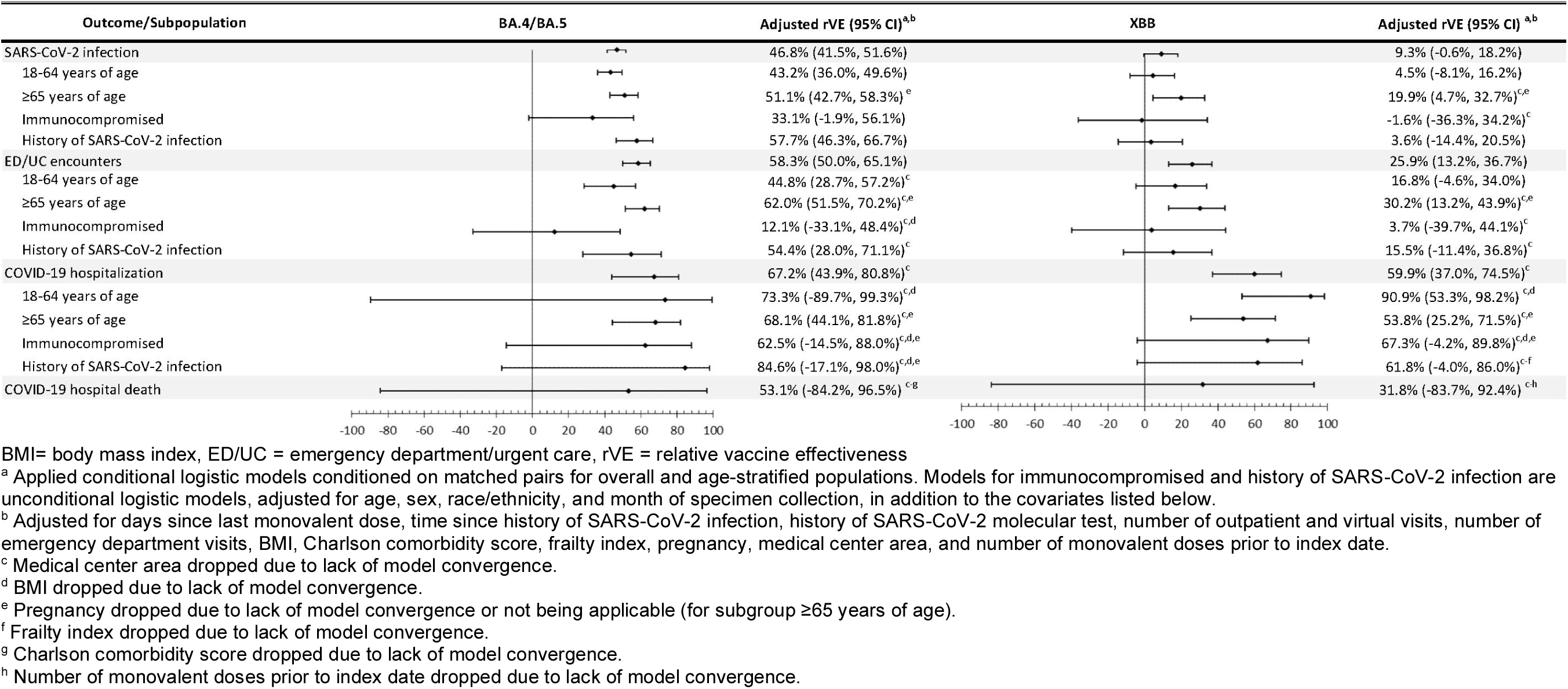
Vaccine effectiveness of mRNA-1273 bivalent vaccine vs. ≥2 monovalent mRNA vaccines against infection and severe outcomes with SARS-CoV-2 variants, overall and by subpopulation

**Figure 4.**
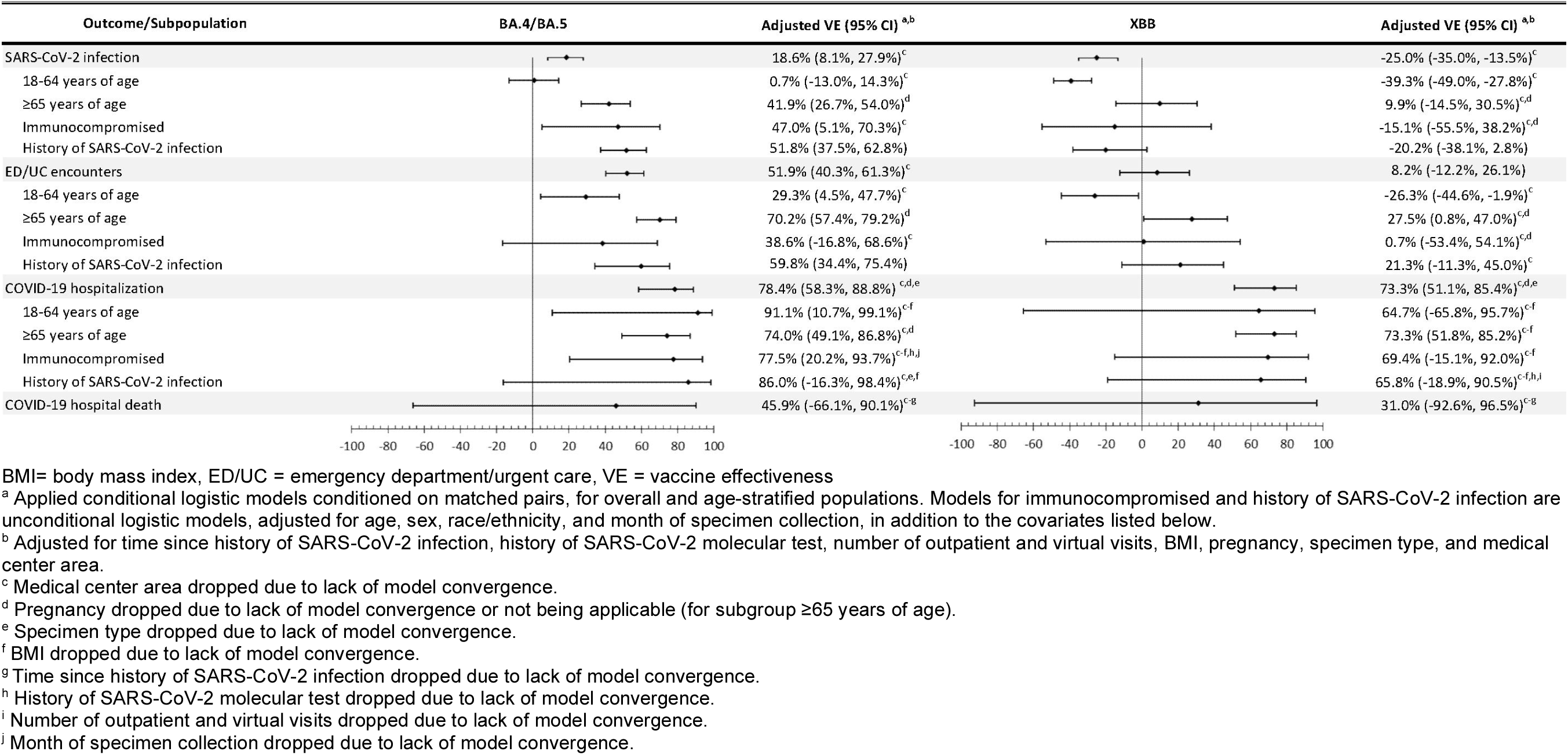
Vaccine effectiveness of mRNA-1273 bivalent vaccine vs. unvaccinated against infection and severe outcomes with SARS-CoV-2 variants, overall and by subpopulation

## Discussion

We evaluated the effectiveness of mRNA-1273 bivalent COVID-19 vaccine against sequencing-confirmed BA.4/BA.5-r and XBB-related sublineages in a large, socio-demographically diverse population. This is the first study to assess long-term bivalent VE against severe outcomes with sequencing-confirmed XBB-related sublineages. We found that rVE against infection and ED/UC encounters with BA.4/BA.5 or XBB was initially similar, but protection against XBB waned and was minimal after 120 days. Although rVE against hospitalization for BA.4/BA.5 or XBB was high initially and waned more slowly than rVE against infection and ED/UC encounters, rVE against hospitalization for XBB declined to 30.0% (-26.0-63.7%) >180 days. These data highlight the need for periodic COVID-19 vaccination as protection wanes against infection and hospitalization, even when vaccine is well-matched to circulating variants.

Furthermore, these findings suggest that periodic adjustment of vaccines to better target emerging variants that can escape vaccine and infection-induced immunity may be beneficial. Data on the effectiveness of BA.4/BA.5 bivalent vaccine against COVID-19 outcomes with XBB-related sublineages are limited. One study in an outpatient pharmacy setting found similar rVE against BA.4/BA.5 and XBB symptomatic infection for up to 3 months [17]. An early study conducted in North Carolina by Lin and colleagues found an initial rVE against hospitalization of 68.3% at 2 weeks that decreased to 30.0% by 8 weeks during an interval when BA.4/BA.5 followed by XBB were predominant; in this study, time since vaccination and the emergence of increasingly evasive sublineages could have contributed to waning [7]. While the estimated rVE against death through 7 weeks in Lin’s study (65.7% [19.7-85.3%]) was somewhat higher than the rVE against death with BA.4/BA.5 or XBB that we observed (53.1% [-84.2-96.5%] and 31.8% [-83.7-92.4%], respectively), our estimates were imprecise and included longer follow-up, potentially reflecting waning, although we were not able to stratify by time. More recently, Link-Gelles and colleagues found lower initial VE against hospitalization during a period that included BA.4/BA.5 and XBB (62% [57-67%] at 7-59 days post-vaccination) compared to our estimated initial VE against hospitalization with BA.4/BA.5 or XBB (76.9% [52.7-88.7%] and 93.4% [68.6-98.6%], respectively, at 14-60 days post-vaccination) and greater waning than our observations, possibly because our analysis was limited to chart-confirmed COVID-19 hospitalizations [8, 18]. rVE and VE against infection with XBB reached statistically significant negative values at >180 and >120 days, respectively. However, negative vaccine effectiveness likely reflects the impact of bias rather than true negative biological effectiveness [19, 20]. Underreporting of prior infection could lead to underestimation of effectiveness and overestimation of waning as the number of controls with natural immunity to SARS-CoV-2 increases over time, biasing effectiveness estimates downward [21, 22]. In addition, increased healthcare-seeking behavior associated with vaccination may result in greater testing and detection of both past and current infection among vaccinees, reducing estimated effectiveness [23]. Furthermore, vaccinated persons may have less infection-avoidant behavior and greater contacts than unvaccinated persons that may reduce estimated effectiveness [24–26]. However, these biases have less impact on estimated effectiveness against more severe outcomes (e.g., hospitalization) for which healthcare seeking behavior and testing vary less by vaccination status.

Like all observational studies, our study has limitations. First, the results of our test-negative case-control study may not be generalizable to people who are not tested, including those with milder symptoms who may not seek testing in healthcare settings. However, the test-negative design reduces bias due to differences in care-seeking behavior since both test-positive cases and test-negative controls have been tested. While there may be residual bias due to factors that were not captured in EHR such as mask use, social distancing, and hygiene practices, we attempted to reduce bias by adjusting for sociodemographic characteristics, prior healthcare utilization, SARS-CoV-2 testing and comorbidities. Second, although rapid antigen test results were included in the history of SARS-CoV-2 infection covariate, at-home positive rapid antigen test results that were not self-reported would be missed. Because both cases and controls had a PCR test performed at KPSC, we expect that the rate of under-reporting of at-home rapid antigen test results would be nondifferential between cases and controls, but it may have differed by vaccination status. Similarly, misclassification of vaccination status was possible but likely minimal as we captured external vaccine administrations from the California Immunization Registry. Third, limited sample size for subgroup analyses and rare outcomes, including COVID-19 hospital death, resulted in wide confidence intervals. Finally, due to the short interval between introduction of bivalent vaccine and the emergence of XBB-sublineages, it was not possible to estimate effectiveness against hospitalization with BA.4/BA.5-related sublineages after 120 days.

This study of mRNA-1273 bivalent vaccine found adequate initial protection against infection with BA.4/BA.5- or XBB-related omicron sublineages that waned quickly against XBB, becoming minimal after 120 days [27]. However, although initially high protection against hospitalization with BA.4/BA.5 or XBB decreased by 120 days, rVE and VE point estimates against XBB hospitalization remained moderately positive after 180 days although with confidence intervals that overlapped zero (30% [-26.0-63.7%] and 42.4% [-19.2-73.2%], respectively). Nonetheless, these data suggest the need for periodic vaccination and adjustment of vaccines to match circulating variants. However, only 17% of the US population received a bivalent COVID-19 vaccine and 4.5% have received the updated XBB.1.5 vaccine as of October 27, 2023.

Consequently, most of the US population is either unvaccinated or has received monovalent vaccine ≥1 year ago, providing negligible protection against infection or severe disease. Greater awareness of disease activity and the effectiveness of updated XBB.1.5 vaccine against COVID-19 outcomes, particularly severe disease, may improve vaccine uptake, potentially reducing COVID-19 associated morbidity and mortality [28].

## Data Availability

Individual-level data reported in this study involving human research participants are not publicly shared due to potentially identifying or sensitive patient information. Upon request to corresponding author [BKA], and subject to review and approval of an analysis proposal, KPSC may provide the deidentified aggregate-level data that support the findings of this study within 6 months. Anonymized data (deidentified data including participant data as applicable) that support the findings of this study may be made available from the investigative team in the following conditions: (1) agreement to collaborate with the study team on all publications, (2) provision of external funding for administrative and investigator time necessary for this collaboration, (3) demonstration that the external investigative team is qualified and has documented evidence of training for human subjects protections, and (4) agreement to abide by the terms outlined in data use agreements between institutions.

## Acknowledgement

The authors would like to acknowledge the following Kaiser Permanente Southern California staff: Maria Navarro, Elsa Olvera, Joy Gelfond, Jonathan Arguello, Diana Romero, Joanna Truong, Samuel Payan, Sierra Lewis, Brittany Brown, and Hiba Atif for their contributions in manual chart reviews of the electronic health records; and Elmer Ayala, Samantha Bayulot, Candice Beissel, Jared Davis, Sarbjit Kaur-Chand, Kourtney Kottmann, Samantha Quinones, Jose Rodriguez, Charanjot (Joe) Singh, Katy Taylor, Joanna Truong, and Vanessa Pan for technical and laboratory support in processing SARS-CoV-2 specimens. The authors would like to acknowledge HelixOpCo, LLC, for their WGS of SARS-CoV-2 specimens. The authors would also like to acknowledge the contributions by Moderna, Inc. staff: Christine Yu and Yamuna Paila. Editorial assistance was provided by Rachel Schultz, MA, ELS, of MEDiSTRAVA in accordance with Good Publication Practice (GPP3) guidelines, funded by Moderna, Inc., and under the direction of the authors. The authors thank the patients of Kaiser Permanente for their partnership with us to improve their health. Their information, collected through our electronic health record systems, leads to findings that help us improve care for our members and can be shared with the larger community.

## Author contributions

BKA, KJB, LQ, LSS, JHK, AF, MAM, CAT, and HFT conceived and designed the study. BKA, KJB, LQ, LSS, SQ, JET, GSL, JHK, AF, YL, RB, JS, SKC, HST, MA, MAM, EJA, CKZ, TS, CAT, and HFT were involved in acquisition, analysis, or interpretation of data. BKA and KJB developed the first draft of the manuscript. LQ, LSS, SQ, JET, GSL, JHK, AF, YL, RB, JS, SKC, HST, MA, MAM, EJA, CKZ, TS, CAT, and HFT were involved in the critical revision of the manuscript for important intellectual content. LQ, YL, JET, and SQ performed the statistical analysis. CAT and HFT obtained funding for the study. GSL, LSS, and MAM provided administrative, technical, or material support. MAM, CAT, and HFT provided project supervision. All authors contributed to the writing of the manuscript and approved the final version for publication.

