## Supplementary Materials for "Effectiveness and durability of mRNA-1273 BA.4/BA.5 bivalent vaccine (mRNA-1273.222) against SARS-CoV-2 BA.4/BA.5 and XBB sublineages"

**SUPPLEMENTARY APPENDIX**

**Authors:** Bradley K. Ackerson[1], Katia J. Bruxvoort[1,3], Lei Qian[1], Lina S. Sy[1], Sijia Qiu[1], Julia E. Tubert[1], Gina S. Lee[1], Jennifer H. Ku[1], Ana Florea[1], Yi Luo[1], Radha Bathala[1], Julie Stern[1], Soon K. Choi[1], Harpreet S. Takhar[1], Michael Aragones[1], Morgan A. Marks[4], Evan J. Anderson[4], Cindy Ke Zhou[4], Tianyu Sun[4], Carla A. Talarico[4,5], Hung Fu Tseng[1,2]

**Affiliations:**

1. Department of Research and Evaluation, Kaiser Permanente Southern California, Pasadena, CA, USA
2. Department of Health Systems Science, Kaiser Permanente Bernard J. Tyson School of Medicine, Pasadena, CA, USA
3. Department of Epidemiology, University of Alabama at Birmingham, Birmingham, AL, USA
4. Moderna Inc., Cambridge, MA, USA
5. AstraZeneca, Gaithersburg, MD, USA

**Corresponding author:**

Bradley Ackerson, MD, Department of Research and Evaluation, Kaiser Permanente Southern California. 100 S. Los Robles Ave, 2nd Floor. Pasadena, CA 91101.

ORCID: 0000-0002-0816-7345

**Alternative corresponding author:**

Hung Fu Tseng, PHD, Department of Research and Evaluation, Kaiser Permanente Southern California. 100 S. Los Robles Ave, 2nd Floor. Pasadena, CA 91101.

ORCID: 0000-0001-6184-6534

Supplementary Table 1. Characteristics of SARS-CoV-2 specimens, by sequencing status and vaccination status

| | Sequencing Success | | | | Sequencing Failure | | | |
|---|---|---|---|---|---|---|---|---|
| | **mRNA-1273 bivalent vaccinated** | **≥2 doses monovalent mRNA** | **Unvaccinated** | **Total** | **mRNA-1273 bivalent vaccinated** | **≥2 doses monovalent mRNA** | **Unvaccinated** | **Total** |
| Overall, n | 1632 | 9946 | 3706 | 15284 | 1445 | 7945 | 3553 | 12943 |
| Specimen type, n (%) | | | | | | | | |
| Nasopharyngeal/oropharyngeal swab | 1477 (90.5%) | 9110 (91.6%) | 3416 (92.2%) | 14003 (91.6%) | 1141 (79.0%) | 6536 (82.3%) | 2900 (81.6%) | 10577 (81.7%) |
| Saliva | 155 (9.5%) | 836 (8.4%) | 290 (7.8%) | 1281 (8.4%) | 304 (21.0%) | 1409 (17.7%) | 653 (18.4%) | 2366 (18.3%) |
| Composite Ct value, n (%)[a] | | | | | | | | |
| ≤27 | 1607 (98.5%) | 9731 (97.8%) | 3619 (97.7%) | 14957 (97.9%) | 285 (19.7%) | 1659 (20.9%) | 651 (18.3%) | 2595 (20.0%) |
| >27 | 25 (1.5%) | 201 (2.0%) | 82 (2.2%) | 308 (2.0%) | 916 (63.4%) | 5058 (63.7%) | 2202 (62.0%) | 8176 (63.2%) |
| Missing | 0 (0.0%) | 14 (0.1%) | 5 (0.1%) | 19 (0.1%) | 244 (16.9%) | 1228 (15.5%) | 700 (19.7%) | 2172 (16.8%) |
| SGTF data available, n (%) | 1212 (74.3%) | 7349 (73.9%) | 2440 (65.8%) | 11001 (72.0%) | 1121 (77.6%) | 6360 (80.1%) | 2740 (77.1%) | 10221 (79.0%) |
| ED/UC encounters | 590 (36.2%) | 3374 (33.9%) | 1444 (39.0%) | 5408 (35.4%) | 504 (34.9%) | 2410 (30.3%) | 1013 (28.5%) | 3927 (30.3%) |
| COVID-19 hospitalization, n (%) | 64 (3.9%) | 427 (4.3%) | 160 (4.3%) | 651 (4.3%) | 55 (3.8%) | 258 (3.2%) | 101 (2.8%) | 414 (3.2%) |
| COVID-19 hospital death, n (%) | 6 (0.4%) | 26 (0.3%) | 8 (0.2%) | 40 (0.3%) | 2 (0.1%) | 10 (0.1%) | 6 (0.2%) | 18 (0.1%) |
| Variant, n (%) | | | | | | | | |
| BA.2 | 72 (4.4%) | 379 (3.8%) | 136 (3.7%) | 587 (3.8%) | N/A | N/A | N/A | N/A |
| BA.4 | 3 (0.2%) | 157 (1.6%) | 70 (1.9%) | 230 (1.5%) | N/A | N/A | N/A | N/A |
| BA.5 (excluding BQ) | 190 (11.6%) | 3020 (30.4%) | 1238 (33.4%) | 4448 (29.1%) | N/A | N/A | N/A | N/A |
| BQ | 558 (34.2%) | 3363 (33.8%) | 1338 (36.1%) | 5259 (34.4%) | N/A | N/A | N/A | N/A |
| XBB.1.5 | 533 (32.7%) | 2045 (20.6%) | 655 (17.7%) | 3233 (21.2%) | N/A | N/A | N/A | N/A |
| XBB.1.16 | 59 (3.6%) | 167 (1.7%) | 39 (1.1%) | 265 (1.7%) | N/A | N/A | N/A | N/A |
| XBB.1.9 | 105 (6.4%) | 297 (3.0%) | 57 (1.5%) | 459 (3.0%) | N/A | N/A | N/A | N/A |
| Other XBB (excluding XBB.1.5, XBB.1.16, XBB.1.9) | 102 (6.3%) | 472 (4.7%) | 147 (4.0%) | 721 (4.7%) | N/A | N/A | N/A | N/A |
| Other | 10 (0.6%) | 46 (0.5%) | 26 (0.7%) | 82 (0.5%) | N/A | N/A | N/A | N/A |

SGTF = S gene target failure, ED/UC = emergency department/urgent care, N/A = not applicable
[a] Cycle threshold (Ct) values from RT-qPCR performed at Helix OpCo, LLC, using a single signal for the combined S and N genes.

Supplementary Table 2. Characteristics of SARS-CoV-2 test-positive cases and test-negative controls with mRNA-1273 bivalent vaccine or unvaccinated

| | Test Positive N=10336 | Test Negative N=31008 | p value | ASD |
|---|---|---|---|---|
| Age at specimen collection date, years | | | 0.42 | 0.01 |
| mean (sd) | 40.46 (24.89) | 40.16 (24.98) | | |
| median | 41 | 40 | | |
| Q1, Q3 | 21, 61 | 20, 61 | | |
| min, max | 0.5, 103 | 0.5, 104 | | |
| Age at specimen collection date, years, n (%) | | | N/A | N/A |
| ≤5 | 1270 (12.3%) | 3810 (12.3%) | | |
| 6-17 | 1118 (10.8%) | 3354 (10.8%) | | |
| 18-44 | 3322 (32.1%) | 9966 (32.1%) | | |
| 45-64 | 2593 (25.1%) | 7779 (25.1%) | | |
| 65-74 | 1119 (10.8%) | 3357 (10.8%) | | |
| ≥75 | 914 (8.8%) | 2742 (8.8%) | | |
| Sex, n (%) | | | N/A | N/A |
| Female | 5462 (52.8%) | 16386 (52.8%) | | |
| Male | 4874 (47.2%) | 14622 (47.2%) | | |
| Race/Ethnicity, n (%) | | | N/A | N/A |
| Non-Hispanic White | 2689 (26.0%) | 8067 (26.0%) | | |
| Non-Hispanic Black | 1058 (10.2%) | 3174 (10.2%) | | |
| Hispanic | 4944 (47.8%) | 14832 (47.8%) | | |
| Non-Hispanic Asian | 1011 (9.8%) | 3033 (9.8%) | | |
| Other/Unknown | 634 (6.1%) | 1902 (6.1%) | | |
| Body mass index[a], $kg/m^2$, n (%) | | | <0.01 | 0.14 |
| <18.5 | 1069 (10.3%) | 3994 (12.9%) | | |
| 18.5 - <25 | 2134 (20.6%) | 6577 (21.2%) | | |
| 25 - <30 | 2414 (23.4%) | 7365 (23.8%) | | |
| 30 - <35 | 1712 (16.6%) | 5320 (17.2%) | | |
| 35 - <40 | 874 (8.5%) | 2657 (8.6%) | | |
| 40 - <45 | 381 (3.7%) | 1108 (3.6%) | | |
| ≥45 | 249 (2.4%) | 800 (2.6%) | | |
| Unknown | 1503 (14.5%) | 3187 (10.3%) | | |
| Smoking[a], n (%) | | | <0.01 | 0.07 |
| No | 7835 (75.8%) | 23471 (75.7%) | | |
| Yes | 1711 (16.6%) | 5642 (18.2%) | | |
| Unknown | 790 (7.6%) | 1895 (6.1%) | | |
| Charlson comorbidity score[b,c] | | | <0.01 | 0.07 |
| mean (sd) | 0.80 (1.69) | 0.92 (1.78) | | |
| median | 0 | 0 | | |
| Q1, Q3 | 0, 1 | 0, 1 | | |
| min, max | 0, 14 | 0, 15 | | |
| Charlson comorbidity score[b,c], n (%) | | | <0.01 | 0.09 |
| 0 | 7078 (68.5%) | 19943 (64.3%) | | |
| 1 | 1541 (14.9%) | 5076 (16.4%) | | |
| ≥2 | 1717 (16.6%) | 5989 (19.3%) | | |

| | | | | |
|---|---|---|---|---|
| Frailty index[b,d] | | | <0.01 | 0.06 |
| mean (sd) | 0.12 (0.03) | 0.12 (0.03) | | |
| median | 0.11 | 0.11 | | |
| Q1, Q3 | 0.10, 0.13 | 0.10, 0.13 | | |
| min, max | 0.06, 0.39 | 0.05, 0.37 | | |
| Frailty index[b,d], n (%) | | | <0.01 | 0.08 |
| Quartile 1 | 2597 (25.1%) | 7741 (25.0%) | | |
| Quartile 2 | 2821 (27.3%) | 7513 (24.2%) | | |
| Quartile 3 | 2505 (24.2%) | 7832 (25.3%) | | |
| Quartile 4, most frail | 2413 (23.3%) | 7922 (25.5%) | | |
| Chronic diseases[b], n (%) | | | | |
| Kidney disease | 630 (6.1%) | 2063 (6.7%) | 0.05 | 0.02 |
| Heart disease | 394 (3.8%) | 1454 (4.7%) | <0.01 | 0.04 |
| Lung disease | 1251 (12.1%) | 4509 (14.5%) | <0.01 | 0.07 |
| Liver disease | 392 (3.8%) | 1401 (4.5%) | <0.01 | 0.04 |
| Diabetes | 1330 (12.9%) | 4255 (13.7%) | 0.03 | 0.03 |
| Immunocompromised, n (%) | | | <0.01 | 0.04 |
| Yes | 421 (4.1%) | 1521 (4.9%) | | |
| HIV/AIDS | 23 | 98 | | |
| Leukemia, lymphoma, congenital and other immunodeficiencies, asplenia/hyposplenia | 176 | 518 | | |
| Organ transplant | 49 | 119 | | |
| Immunosuppressant medications | 272 | 1031 | | |
| Autoimmune conditions[b], n (%) | | | 0.94 | <0.01 |
| Yes | 336 (3.3%) | 1013 (3.3%) | | |
| Rheumatoid arthritis | 140 | 451 | | |
| Inflammatory bowel disease | 65 | 234 | | |
| Psoriasis and psoriatic arthritis | 123 | 313 | | |
| Multiple sclerosis | 25 | 47 | | |
| Systemic lupus erythematosus | 32 | 102 | | |
| Pregnant at specimen collection date, n (%) | | | <0.01 | 0.16 |
| Yes | 198 (1.9%) | 1453 (4.7%) | | |
| 1st trimester | 28 | 100 | | |
| 2nd trimester | 53 | 129 | | |
| 3rd trimester | 117 | 1224 | | |
| History of SARS-CoV-2 infection[e], n (%) | | | <0.01 | 0.11 |
| Yes | 3222 (31.2%) | 11215 (36.2%) | | |
| <180 days | 173 | 1652 | | |
| 180- <360 days | 1292 | 4436 | | |
| ≥360 days | 1757 | 5127 | | |
| History of SARS-CoV-2 molecular test[e], n (%) | 8131 (78.7%) | 22539 (72.7%) | <0.01 | 0.14 |
| Number of outpatient and virtual visits[b], n (%) | | | <0.01 | 0.11 |
| 0 | 732 (7.1%) | 1594 (5.1%) | | |
| 1-4 | 2672 (25.9%) | 7649 (24.7%) | | |
| 5-10 | 2966 (28.7%) | 8474 (27.3%) | | |
| ≥11 | 3966 (38.4%) | 13291 (42.9%) | | |
| Number of Emergency Department visits[b], n (%) | | | <0.01 | 0.10 |
| 0 | 7784 (75.3%) | 21943 (70.8%) | | |
| 1 | 1583 (15.3%) | 5759 (18.6%) | | |

| | | | | |
|---|---|---|---|---|
| ≥2 | 969 (9.4%) | 3306 (10.7%) | | |
| Number of hospitalizations[b], n (%) | | | <0.01 | 0.05 |
| 0 | 9383 (90.8%) | 28406 (91.6%) | | |
| 1 | 764 (7.4%) | 1958 (6.3%) | | |
| ≥2 | 189 (1.8%) | 644 (2.1%) | | |
| Preventive care[b], n (%) | 7434 (71.9%) | 22607 (72.9%) | 0.05 | 0.02 |
| Medicaid, n (%) | 1592 (15.4%) | 5894 (19.0%) | <0.01 | 0.10 |
| Neighborhood median household income, n (%) | | | 0.02 | 0.04 |
| < $40,000 | 180 (1.7%) | 603 (1.9%) | | |
| $40,000-$59,999 | 1728 (16.7%) | 5037 (16.2%) | | |
| $60,000-$79,999 | 2602 (25.2%) | 7597 (24.5%) | | |
| ≥$80,000 | 5781 (55.9%) | 17684 (57.0%) | | |
| Unknown | 45 (0.4%) | 87 (0.3%) | | |
| Medical center area[f] | | | <0.01 | 0.17 |
| Month of specimen collection, n (%) | | | <0.01 | 0.07 |
| September - October 2022 | 1590 (15.4%) | 5101 (16.5%) | | |
| November - December 2022 | 4582 (44.3%) | 12771 (41.2%) | | |
| January - February 2023 | 2186 (21.1%) | 7033 (22.7%) | | |
| March - April 2023 | 1064 (10.3%) | 3395 (10.9%) | | |
| May - June 2023 | 914 (8.8%) | 2708 (8.7%) | | |
| Specimen type, n (%) | | | <0.01 | 0.15 |
| Nasopharyngeal/oropharyngeal swab | 8934 (86.4%) | 28224 (91.0%) | | |
| Saliva | 1402 (13.6%) | 2784 (9.0%) | | |
| Vaccination status, n (%) | | | 0.74 | <0.01 |
| Bivalent vaccinated | 3077 (29.8%) | 9285 (29.9%) | | |
| Unvaccinated | 7259 (70.2%) | 21723 (70.1%) | | |
| Antiviral therapy within 7 days after the index date n (%) | | | N/A | N/A |
| Yes | 1372 (13.3%) | N/A | N/A | N/A |
| Nirmatrelvir/ritonavir | 1352 | N/A | N/A | N/A |
| Molnupiravir | 6 | N/A | N/A | N/A |
| Remdesivir | 14 | N/A | N/A | N/A |

ASD = absolute standardized difference, sd = standard deviation, N/A = not applicable
[a] Defined in the two years prior to specimen collection date
[b] Defined in the one year prior to specimen collection date
[c] Possible range: 0-29. Quan, H., et al. Updating and validating the Charlson comorbidity index and score for risk adjustment in hospital discharge abstracts using data from 6 countries. American Journal of Epidemiology 173.6 (2011): 676-682.
[d] Possible range: 0-1. Kim, D.H., et al. Measuring frailty in Medicare data: development and validation of a claims-based frailty index. The Journals of Gerontology: Series A 73.7 (2018): 980-987.
[e] Defined based on all available medical records from March 1, 2020 to specimen collection date
[f] Frequency and percent for the 19 medical center areas not shown

Supplementary Table 3. Vaccine effectiveness of mRNA-1273 bivalent vaccine vs. ≥2 monovalent mRNA vaccines against infection and severe outcomes with SARS-CoV-2 variants by time since vaccination

| Subvariant/Time since vaccination | Test Positive | | Test Negative | | Odds Ratio (95% CI) | | rVE (95% CI)[a] | |
|---|---|---|---|---|---|---|---|---|
| | mRNA-1273 bivalent vaccinated (%) | ≥2 doses monovalent mRNA vaccinated (%) | mRNA-1273 bivalent vaccinated (%) | ≥2 doses monovalent mRNA vaccinated (%) | Unadjusted | Adjusted [b,c] | Unadjusted | Adjusted [b,c] |
| BA.4/BA.5 | | | | | | | | |
| SARS-CoV-2 infection | 751 (10.3%) | 6540 (89.7%) | 3373 (15.4%) | 18500 (84.6%) | 0.597 (0.547, 0.652) | 0.532 (0.484, 0.585) | 40.3% (34.8%, 45.3%) | 46.8% (41.5%, 51.6%) |
| 14-60 days | 432 (6.2%) | 6540 (93.8%) | 2186 (10.6%) | 18500 (89.4%) | 0.559 (0.502, 0.622) | 0.473 (0.422, 0.531) | 44.1% (37.8%, 49.8%) | 52.7% (46.9%, 57.8%) |
| 61-120 days | 293 (4.3%) | 6540 (95.7%) | 1096 (5.6%) | 18500 (94.4%) | 0.756 (0.663, 0.863) | 0.606 (0.525, 0.699) | 24.4% (13.7%, 33.7%) | 39.4% (30.1%, 47.5%) |
| 121-180 days | 25 (0.4%) | 6540 (99.6%) | 91 (0.5%) | 18500 (99.5%) | 0.777 (0.499, 1.211) | 0.645 (0.405, 1.029) | 22.3% (-17.4%, 50.1%) | 35.5% (-2.8%, 59.5%) |
| >180 days | 1 (0.0%) | 6540 (100.0%) | 0 (0.0%) | 18500 (100.0%) | N/A | N/A | N/A | N/A |
| ED/UC encounters | 218 (10.1%) | 1930 (89.9%) | 1190 (18.5%) | 5254 (81.5%) | 0.461 (0.392, 0.542) | 0.417 (0.349, 0.500) | 53.9% (45.8%, 60.8%) | 58.3% (50.0%, 65.1%) |
| 14-60 days [d] | 125 (6.1%) | 1930 (93.9%) | 743 (12.4%) | 5254 (87.6%) | 0.458 (0.376, 0.557) | 0.407 (0.330, 0.503) | 54.2% (44.3%, 62.4%) | 59.3% (49.7%, 67.0%) |
| 61-120 days [d] | 82 (4.1%) | 1930 (95.9%) | 410 (7.2%) | 5254 (92.8%) | 0.544 (0.427, 0.694) | 0.490 (0.376, 0.637) | 45.6% (30.6%, 57.3%) | 51.0% (36.3%, 62.4%) |
| 121-180 days [d] | 10 (0.5%) | 1930 (99.5%) | 37 (0.7%) | 5254 (99.3%) | 0.736 (0.365, 1.482) | 0.668 (0.320, 1.393) | 26.4% (-32.5%, 63.5%) | 33.2% (-28.2%, 68.0%) |
| >180 days [d] | 1 (0.1%) | 1930 (99.9%) | 0 (0.0%) | 5254 (100.0%) | N/A | N/A | N/A | N/A |
| COVID-19 hospitalization [d] | 24 (9.3%) | 235 (90.7%) | 196 (25.2%) | 581 (74.8%) | 0.289 (0.183, 0.456) | 0.328 (0.192, 0.561) | 71.1% (54.4%, 81.7%) | 67.2% (43.9%, 80.8%) |
| 14-60 days [d] | 12 (4.9%) | 235 (95.1%) | 129 (18.2%) | 581 (81.8%) | 0.230 (0.125, 0.424) | 0.287 (0.149, 0.551) | 77.0% (57.6%, 87.5%) | 71.3% (44.9%, 85.1%) |
| 61-120 days [d] | 11 (4.5%) | 235 (95.5%) | 65 (10.1%) | 581 (89.9%) | 0.418 (0.217, 0.807) | 0.480 (0.227, 1.012) | 58.2% (19.3%, 78.3%) | 52.0% (-1.2%, 77.3%) |
| 121-180 days [d] | 0 (0.0%) | 235 (100.0%) | 2 (0.3%) | 581 (99.7%) | N/A | N/A | N/A | N/A |
| >180 days [d] | 1 (0.4%) | 235 (99.6%) | 0 (0.0%) | 581 (100.0%) | N/A | N/A | N/A | N/A |
| COVID-19 hospital death [d,e] | 2 (11.8%) | 15 (88.2%) | 13 (25.5%) | 38 (74.5%) | 0.358 (0.069, 1.869) | 0.469 (0.035, 6.337) | 64.2% (-46.5%, 93.1%) | 53.1% (-84.2%, 96.5%) |
| XBB | | | | | | | | |
| SARS-CoV-2 infection | 799 (21.1%) | 2981 (78.9%) | 2240 (19.8%) | 9100 (80.2%) | 1.095 (0.997, 1.203) | 0.907 (0.818, 1.006) | -8.7% (-16.9%, 0.3%) | 9.3% (-0.6%, 18.2%) |
| 14-60 days | 76 (2.5%) | 2981 (97.5%) | 356 (3.8%) | 9100 (96.2%) | 0.652 (0.507, 0.838) | 0.512 (0.393, 0.666) | 34.8% (16.2%, 49.3%) | 48.8% (33.4%, 60.7%) |
| 61-120 days | 212 (6.6%) | 2981 (93.4%) | 698 (7.1%) | 9100 (92.9%) | 0.927 (0.791, 1.087) | 0.740 (0.623, 0.879) | 7.3% (-8.0%, 20.9%) | 26.0% (12.1%, 37.7%) |
| 121-180 days | 265 (8.2%) | 2981 (91.8%) | 662 (6.8%) | 9100 (93.2%) | 1.222 (1.054, 1.417) | 1.041 (0.887, 1.221) | -18.2% (-29.5%, -5.1%) | -3.9% (-18.1%, 11.3%) |
| >180 days | 246 (7.6%) | 2981 (92.4%) | 524 (5.4%) | 9100 (94.6%) | 1.433 (1.225, 1.677) | 1.225 (1.030, 1.456) | -30.2% (-40.4%, -18.4%) | -18.3% (-31.3%, -2.9%) |

| | | | | | | | | |
|---|---|---|---|---|---|---|---|---|
| ED/UC encounters | 343 (20.8%) | 1307 (79.2%) | 1113 (22.5%) | 3837 (77.5%) | 0.899 (0.782, 1.035) | 0.741 (0.633, 0.868) | 10.1% (-3.4%, 21.8%) | 25.9% (13.2%, 36.7%) |
| 14-60 days [d] | 20 (1.5%) | 1307 (98.5%) | 158 (4.0%) | 3837 (96.0%) | 0.372 (0.232, 0.594) | 0.293 (0.180, 0.476) | 62.8% (40.6%, 76.8%) | 70.7% (52.4%, 82.0%) |
| 61-120 days [d] | 92 (6.6%) | 1307 (93.4%) | 304 (7.3%) | 3837 (92.7%) | 0.888 (0.698, 1.131) | 0.697 (0.537, 0.905) | 11.2% (-11.6%, 30.2%) | 30.3% (9.5%, 46.3%) |
| 121-180 days [d] | 118 (8.3%) | 1307 (91.7%) | 357 (8.5%) | 3837 (91.5%) | 0.970 (0.781, 1.206) | 0.843 (0.668, 1.063) | 3.0% (-17.1%, 21.9%) | 15.7% (-6.0%, 33.2%) |
| >180 days [d] | 113 (8.0%) | 1307 (92.0%) | 294 (7.1%) | 3837 (92.9%) | 1.128 (0.900, 1.414) | 0.980 (0.764, 1.257) | -11.4% (-29.3%, 10.0%) | 2.0% (-20.4%, 23.6%) |
| COVID-19 hospitalization [d] | 40 (18.9%) | 172 (81.1%) | 209 (32.9%) | 427 (67.1%) | 0.482 (0.329, 0.705) | 0.401 (0.255, 0.630) | 51.8% (29.5%, 67.1%) | 59.9% (37.0%, 74.5%) |
| 14-60 days [d] | 2 (1.1%) | 172 (98.9%) | 27 (5.9%) | 427 (94.1%) | 0.184 (0.043, 0.782) | 0.121 (0.026, 0.562) | 81.6% (21.8%, 95.7%) | 87.9% (43.8%, 97.4%) |
| 61-120 days [d] | 7 (3.9%) | 172 (96.1%) | 54 (11.2%) | 427 (88.8%) | 0.322 (0.144, 0.721) | 0.219 (0.090, 0.529) | 67.8% (27.9%, 85.6%) | 78.1% (47.1%, 91.0%) |
| 121-180 days [d] | 14 (7.5%) | 172 (92.5%) | 72 (14.4%) | 427 (85.6%) | 0.483 (0.265, 0.879) | 0.429 (0.222, 0.830) | 51.7% (12.1%, 73.5%) | 57.1% (17.0%, 77.8%) |
| >180 days [d] | 17 (9.0%) | 172 (91.0%) | 56 (11.6%) | 427 (88.4%) | 0.754 (0.426, 1.334) | 0.700 (0.363, 1.351) | 24.6% (-25.0%, 57.4%) | 30.0% (-26.0%, 63.7%) |
| COVID-19 hospital death [d,e,f] | 4 (28.6%) | 10 (71.4%) | 17 (40.5%) | 25 (59.5%) | 0.602 (0.166, 2.174) | 0.682 (0.076, 6.133) | 39.8% (-54.0%, 83.4%) | 31.8% (-83.7%, 92.4%) |

BMI= body mass index, ED/UC = emergency department/urgent care, OR = odds ratio, rVE = relative vaccine effectiveness, N/A = not applicable

[a] Calculated as (1 – OR) x 100 when OR was ≤1, and ([1/OR] – 1) × 100 when OR was >1.

[b] Applied conditional logistic models conditioned on matched pairs for SARS-CoV-2 infection, ED/UC encounters, hospitalization and hospital death outcomes. Models for time since vaccination analyses are unconditional logistic models, adjusted for age, sex, race/ethnicity, and month of specimen collection, in addition to the covariates listed below.

[c] Adjusted for days since last monovalent dose, time since history of SARS-CoV-2 infection, history of SARS-CoV-2 molecular test, number of outpatient and virtual visits, number of emergency department visits, BMI, Charlson comorbidity score, frailty index, pregnancy, medical center area, and number of monovalent doses prior to index date.

[d] Medical center area dropped due to lack of model convergence.

[e] BMI, Charlson comorbidity score, frailty index, and pregnancy dropped due to lack of model convergence.

[f] Number of monovalent doses prior to index date dropped due to lack of model convergence.

Supplementary Table 4. Vaccine effectiveness of mRNA-1273 bivalent vaccine vs. unvaccinated against infection and severe outcomes with SARS-CoV-2 variants by time since vaccination

| Subvariant/Time since vaccination | Test Positive | | Test Negative | | Odds Ratio (95% CI) | | VE (95% CI)[a] | |
|---|---|---|---|---|---|---|---|---|
| | mRNA-1273 bivalent vaccinated (%) | Unvaccinated (%) | mRNA-1273 bivalent vaccinated (%) | Unvaccinated (%) | Unadjusted | Adjusted [b,c] | Unadjusted | Adjusted [b,c] |
| BA.4/BA.5 | | | | | | | | |
| SARS-CoV-2 infection [d] | 751 (22.1%) | 2646 (77.9%) | 2478 (24.3%) | 7713 (75.7%) | 0.825 (0.735, 0.925) | 0.814 (0.721, 0.919) | 17.5% (7.5%, 26.5%) | 18.6% (8.1%, 27.9%) |
| 14-60 days [d] | 432 (14.0%) | 2646 (86.0%) | 1615 (17.3%) | 7713 (82.7%) | 0.780 (0.695, 0.875) | 0.707 (0.618, 0.809) | 22.0% (12.5%, 30.5%) | 29.3% (19.1%, 38.2%) |
| 61-120 days [d] | 293 (10.0%) | 2646 (90.0%) | 791 (9.3%) | 7713 (90.7%) | 1.080 (0.938, 1.243) | 0.972 (0.826, 1.143) | -7.4% (-19.6%, 6.2%) | 2.8% (-12.5%, 17.4%) |
| 121-180 days [d] | 25 (0.9%) | 2646 (99.1%) | 70 (0.9%) | 7713 (99.1%) | 1.041 (0.658, 1.647) | 1.097 (0.670, 1.796) | -3.9% (-39.3%, 34.2%) | -8.8% (-44.3%, 33.0%) |
| >180 days [d] | 1 (0.0%) | 2646 (100.0%) | 2 (0.0%) | 7713 (100.0%) | 1.458 (0.132, 16.08) | 1.241 (0.078, 19.75) | -31.4% (-93.8%, 86.8%) | -19.4% (-94.9%, 92.2%) |
| ED/UC encounters[d] | 218 (19.4%) | 905 (80.6%) | 920 (27.3%) | 2449 (72.7%) | 0.507 (0.413, 0.622) | 0.481 (0.387, 0.597) | 49.3% (37.8%, 58.7%) | 51.9% (40.3%, 61.3%) |
| 14-60 days [d] | 125 (12.1%) | 905 (87.9%) | 592 (19.5%) | 2449 (80.5%) | 0.571 (0.464, 0.703) | 0.444 (0.349, 0.566) | 42.9% (29.7%, 53.6%) | 55.6% (43.4%, 65.1%) |
| 61-120 days [d] | 82 (8.3%) | 905 (91.7%) | 302 (11.0%) | 2449 (89.0%) | 0.735 (0.569, 0.949) | 0.581 (0.435, 0.778) | 26.5% (5.1%, 43.1%) | 41.9% (22.2%, 56.5%) |
| 121-180 days [d] | 10 (1.1%) | 905 (98.9%) | 24 (1.0%) | 2449 (99.0%) | 1.128 (0.537, 2.367) | 1.055 (0.473, 2.353) | -11.3% (-57.8%, 46.3%) | -5.2% (-57.5%, 52.7%) |
| >180 days [d] | 1 (0.1%) | 905 (99.9%) | 2 (0.1%) | 2449 (99.9%) | 1.353 (0.123, 14.94) | 1.448 (0.082, 25.70) | -26.1% (-93.3%, 87.7%) | -30.9% (-96.1%, 91.8%) |
| COVID-19 hospitalization [d,e] | 24 (19.8%) | 97 (80.2%) | 173 (47.7%) | 190 (52.3%) | 0.211 (0.121, 0.368) | 0.216 (0.112, 0.417) | 78.9% (63.2%, 87.9%) | 78.4% (58.3%, 88.8%) |
| 14-60 days [d,e] | 12 (11.0%) | 97 (89.0%) | 101 (34.7%) | 190 (65.3%) | 0.233 (0.122, 0.444) | 0.231 (0.113, 0.473) | 76.7% (55.6%, 87.8%) | 76.9% (52.7%, 88.7%) |
| 61-120 days [d,e] | 11 (10.2%) | 97 (89.8%) | 67 (26.1%) | 190 (73.9%) | 0.322 (0.162, 0.637) | 0.321 (0.152, 0.679) | 67.8% (36.3%, 83.8%) | 67.9% (32.1%, 84.8%) |
| 121-180 days [d,e] | 0 (0.0%) | 97 (100.0%) | 5 (2.6%) | 190 (97.4%) | N/A | N/A | N/A | N/A |
| >180 days [d,e] | 1 (1.0%) | 97 (99.0%) | 0 (0.0%) | 190 (100.0%) | N/A | N/A | N/A | N/A |
| COVID-19 hospital death [d,e,f] | 2 (22.2%) | 7 (77.8%) | 10 (37.0%) | 17 (63.0%) | 0.518 (0.097, 2.755) | 0.541 (0.099, 2.947) | 48.2% (-63.7%, 90.3%) | 45.9% (-66.1%, 90.1%) |
| XBB | | | | | | | | |
| SARS-CoV-2 infection [d] | 799 (47.1%) | 898 (52.9%) | 2123 (41.7%) | 2968 (58.3%) | 1.377 (1.203, 1.576) | 1.334 (1.156, 1.538) | -27.4% (-36.5%, -16.9%) | -25.0% (-35.0%, -13.5%) |
| 14-60 days [d] | 76 (7.8%) | 898 (92.2%) | 308 (9.4%) | 2968 (90.6%) | 0.816 (0.628, 1.060) | 0.788 (0.593, 1.048) | 18.4% (-5.6%, 37.2%) | 21.2% (-4.6%, 40.7%) |
| 61-120 days [d] | 212 (19.1%) | 898 (80.9%) | 620 (17.3%) | 2968 (82.7%) | 1.130 (0.951, 1.343) | 1.154 (0.948, 1.406) | -11.5% (-25.6%, 4.9%) | -13.4% (-28.9%, 5.2%) |

| | | | | | | | | |
|---|---|---|---|---|---|---|---|---|
| 121-180 days [d] | 265 (22.8%) | 898 (77.2%) | 683 (18.7%) | 2968 (81.3%) | 1.282 (1.092, 1.505) | 1.429 (1.184, 1.724) | -22.0% (-33.6%, -8.5%) | -30.0% (-42.0%, -15.5%) |
| >180 days [d] | 246 (21.5%) | 898 (78.5%) | 512 (14.7%) | 2968 (85.3%) | 1.588 (1.341, 1.881) | 1.948 (1.578, 2.405) | -37.0% (-46.8%, -25.4%) | -48.7% (-58.4%, -36.6%) |
| ED/UC encounters | 343 (42.1%) | 472 (57.9%) | 1032 (42.2%) | 1413 (57.8%) | 0.992 (0.815, 1.209) | 0.918 (0.739, 1.139) | 0.8% (-17.3%, 18.5%) | 8.2% (-12.2%, 26.1%) |
| 14-60 days | 20 (4.1%) | 472 (95.9%) | 139 (9.0%) | 1413 (91.0%) | Tab | 0.396 (0.237, 0.662) | 56.9% (30.4%, 73.3%) | 60.4% (33.8%, 76.3%) |
| 61-120 days | 92 (16.3%) | 472 (83.7%) | 278 (16.4%) | 1413 (83.6%) | 0.991 (0.766, 1.282) | 0.903 (0.674, 1.210) | 0.9% (-22.0%, 23.4%) | 9.7% (-17.3%, 32.6%) |
| 121-180 days | 118 (20.0%) | 472 (80.0%) | 341 (19.4%) | 1413 (80.6%) | 1.036 (0.820, 1.309) | 0.978 (0.740, 1.292) | -3.5% (-23.6%, 18.0%) | 2.2% (-22.6%, 26.0%) |
| >180 days | 113 (19.3%) | 472 (80.7%) | 274 (16.2%) | 1413 (83.8%) | 1.235 (0.969, 1.574) | 1.172 (0.874, 1.570) | -19.0% (-36.5%, 3.1%) | -14.6% (-36.3%, 12.6%) |
| COVID-19 hospitalization [d,e] | 40 (43.0%) | 53 (57.0%) | 194 (69.5%) | 85 (30.5%) | 0.287 (0.170, 0.486) | 0.267 (0.146, 0.488) | 71.3% (51.4%, 83.0%) | 73.3% (51.2%, 85.4%) |
| 14-60 days [d,e] | 2 (3.6%) | 53 (96.4%) | 35 (29.2%) | 85 (70.8%) | 0.092 (0.021, 0.397) | 0.066 (0.014, 0.314) | 90.8% (60.3%, 97.9%) | 93.4% (68.6%, 98.6%) |
| 61-120 days [d,e] | 7 (11.7%) | 53 (88.3%) | 58 (40.6%) | 85 (59.4%) | 0.194 (0.082, 0.456) | 0.146 (0.056, 0.380) | 80.6% (54.4%, 91.8%) | 85.4% (62.0%, 94.4%) |
| 121-180 days [d,e] | 14 (20.9%) | 53 (79.1%) | 55 (39.3%) | 85 (60.7%) | 0.408 (0.207, 0.806) | 0.362 (0.164, 0.799) | 59.2% (19.4%, 79.3%) | 63.8% (20.1%, 83.6%) |
| >180 days [d,e] | 17 (24.3%) | 53 (75.7%) | 46 (35.1%) | 85 (64.9%) | 0.593 (0.308, 1.139) | 0.576 (0.268, 1.237) | 40.7% (-12.2%, 69.2%) | 42.4% (-19.2%, 73.2%) |
| COVID-19 hospital death[d,e,f] | 4 (80.0%) | 1 (20.0%) | 13 (86.7%) | 2 (13.3%) | 0.577 (0.033, 10.25) | 0.690 (0.035, 13.55) | 42.3% (-90.2%, 96.7%) | 31.0% (-92.6%, 96.5%) |

BMI= body mass index, ED/UC = emergency department/urgent care, OR = odds ratio, VE = vaccine effectiveness, N/A = not applicable

[a] Calculated as (1 – OR) x 100 when OR was ≤1, and ([1/OR] – 1) × 100 when OR was >1.

[b] Applied conditional logistic models conditioned on matched pairs for SARS-CoV-2 infection, ED/UC encounters, hospitalization and hospital death outcomes. Models for time since vaccination analyses are unconditional logistic models, adjusted for age, sex, race/ethnicity, and month of specimen collection, in addition to the covariates listed below.

[c] Adjusted for time since history of SARS-CoV-2 infection, history of SARS-CoV-2 molecular test, number of outpatient and virtual visits, BMI, pregnancy, specimen type, and medical center area.

[d] Medical center area dropped due to lack of model convergence.

[e] Specimen type and pregnancy dropped due to lack of model convergence.

[f] BMI and time since history of SARS-CoV-2 infection dropped due to lack of model convergence.

Supplementary Table 5. Vaccine effectiveness of mRNA-1273 bivalent vaccine vs. ≥2 monovalent mRNA vaccines against infection and severe outcomes with SARS-CoV-2 variants by age group

| | Test Positive | | Test Negative | | Odds Ratio (95% CI) | | rVE (95% CI)[a] | |
|---|---|---|---|---|---|---|---|---|
| **Subvariant/Age group/Outcome** | **mRNA-1273 bivalent vaccinated (%)** | **≥2 doses monovalent mRNA vaccinated (%)** | **mRNA-1273 bivalent vaccinated (%)** | **≥2 doses monovalent mRNA vaccinated (%)** | **Unadjusted** | **Adjusted[b,c]** | **Unadjusted** | **Adjusted[b,c]** |
| BA.4/BA.5 | | | | | | | | |
| Aged ≤17 years | | | | | | | | |
| SARS-CoV-2 infection [d,e] | 4 (9.5%) | 38 (90.5%) | 10 (7.9%) | 116 (92.1%) | 1.591 (0.254, 9.950) | 1.518 (0.240, 9.605) | -37.1% (-90.0%, 74.6%) | -34.1% (-89.6%, 76.0%) |
| ED/UC encounters | 2 (14.3%) | 12 (85.7%) | 7 (16.7%) | 35 (83.3%) | N/A | N/A | N/A | N/A |
| COVID-19 hospitalization | 0 | 0 | 0 | 0 | N/A | N/A | N/A | N/A |
| COVID-19 hospital death | 0 | 0 | 0 | 0 | N/A | N/A | N/A | N/A |
| Aged 18-64 years | | | | | | | | |
| SARS-CoV-2 infection | 446 (8.4%) | 4878 (91.6%) | 1928 (12.1%) | 14044 (87.9%) | 0.647 (0.579, 0.724) | 0.568 (0.504, 0.640) | 35.3% (27.6%, 42.1%) | 43.2% (36.0%, 49.6%) |
| ED/UC encounters [d] | 100 (8.4%) | 1086 (91.6%) | 441 (12.4%) | 3117 (87.6%) | 0.631 (0.499, 0.799) | 0.552 (0.428, 0.713) | 36.9% (20.1%, 50.1%) | 44.8% (28.7%, 57.2%) |
| COVID-19 hospitalization [d,f] | 1 (2.9%) | 33 (97.1%) | 11 (10.8%) | 91 (89.2%) | 0.273 (0.035, 2.112) | 0.267 (0.007, 9.674) | 72.7% (-52.7%, 96.5%) | 73.3% (-89.7%, 99.3%) |
| COVID-19 hospital death | 0 (0.0%) | 3 (100.0%) | 0 (0.0%) | 9 (100.0%) | N/A | N/A | N/A | N/A |
| Aged ≥65 years | | | | | | | | |
| SARS-CoV-2 infection | 301 (15.6%) | 1624 (84.4%) | 1435 (24.8%) | 4340 (75.2%) | 0.522 (0.452, 0.603) | 0.489 (0.417, 0.573) | 47.8% (39.7%, 54.8%) | 51.1% (42.7%, 58.3%) |
| ED/UC encounters[d] | 116 (12.2%) | 832 (87.8%) | 742 (26.1%) | 2102 (73.9%) | 0.362 (0.290, 0.451) | 0.380 (0.298, 0.485) | 63.8% (54.9%, 71.0%) | 62.0% (51.5%, 70.2%) |
| COVID-19 hospitalization [d] | 23 (10.2%) | 202 (89.8%) | 185 (27.4%) | 490 (72.6%) | 0.290 (0.181, 0.463) | 0.319 (0.182, 0.559) | 71.0% (53.7%, 81.9%) | 68.1% (44.1%, 81.8%) |
| COVID-19 hospital death [d-g] | 2 (14.3%) | 12 (85.7%) | 13 (31.0%) | 29 (69.0%) | 0.358 (0.069, 1.869) | 0.257 (0.022, 3.062) | 64.2% (-46.5%, 93.1%) | 74.3% (-67.3%, 97.8%) |
| XBB | | | | | | | | |
| Aged ≤17 years | | | | | | | | |
| SARS-CoV-2 infection | 4 (3.2%) | 121 (96.8%) | 5 (1.3%) | 370 (98.7%) | 2.637 (0.646, 10.76) | 3.691 (0.809, 16.84) | -62.1% (-90.7%, 35.4%) | -72.9% (-94.1%, 19.1%) |
| ED/UC encounters | 0 (0.0%) | 53 (100.0%) | 4 (2.5%) | 155 (97.5%) | N/A | N/A | N/A | N/A |
| COVID-19 hospitalization | 0 | 0 | 0 | 0 | N/A | N/A | N/A | N/A |
| COVID-19 hospital death | 0 | 0 | 0 | 0 | N/A | N/A | N/A | N/A |
| Aged 18-64 years | | | | | | | | |
| SARS-CoV-2 infection | 454 (17.1%) | 2199 (82.9%) | 1224 (15.4%) | 6735 (84.6%) | 1.140 (1.012, 1.285) | 0.968 (0.851, 1.102) | -12.3% (-22.2%, -1.1%) | 4.5% (-8.1%, 16.2%) |

| | | | | | | | | |
|---|---|---|---|---|---|---|---|---|
| ED/UC encounters | 150 (16.0%) | 786 (84.0%) | 445 (15.8%) | 2363 (84.2%) | 1.014 (0.826, 1.245) | 0.845 (0.671, 1.063) | -1.4% (-19.7%, 17.4%) | 16.8% (-4.6%, 34.0%) |
| COVID-19 hospitalization [d,f] | 4 (9.5%) | 38 (90.5%) | 36 (28.6%) | 90 (71.4%) | 0.268 (0.090, 0.800) | 0.091 (0.018, 0.467) | 73.2% (20.0%, 91.0%) | 90.9% (53.3%, 98.2%) |
| COVID-19 hospital death | 1 (50.0%) | 1 (50.0%) | 2 (33.3%) | 4 (66.7%) | 1.732 (0.098, 30.76) | N/A | -42.3% (-96.7%, 90.2%) | N/A |
| Aged ≥65 years | | | | | | | | |
| SARS-CoV-2 infection [d] | 341 (34.0%) | 661 (66.0%) | 1011 (33.6%) | 1995 (66.4%) | 1.018 (0.875, 1.184) | 0.801 (0.673, 0.953) | -1.8% (-15.6%, 12.5%) | 19.9% (4.7%, 32.7%) |
| ED/UC encounters [d] | 193 (29.2%) | 468 (70.8%) | 664 (33.5%) | 1319 (66.5%) | 0.820 (0.677, 0.993) | 0.698 (0.561, 0.868) | 18.0% (0.7%, 32.3%) | 30.2% (13.2%, 43.9%) |
| COVID-19 hospitalization [d] | 36 (21.2%) | 134 (78.8%) | 173 (33.9%) | 337 (66.1%) | 0.532 (0.353, 0.801) | 0.462 (0.285, 0.748) | 46.8% (19.9%, 64.7%) | 53.8% (25.2%, 71.5%) |
| COVID-19 hospital death [d-g] | 3 (25.0%) | 9 (75.0%) | 15 (41.7%) | 21 (58.3%) | 0.472 (0.111, 2.014) | 0.248 (0.018, 3.430) | 52.8% (-50.3%, 88.9%) | 75.2% (-70.8%, 98.2%) |

BMI= body mass index, ED/UC = emergency department/urgent care, OR = odds ratio, rVE = relative vaccine effectiveness, N/A = not applicable

[a] Calculated as (1 – OR) x 100 when OR was ≤1, and ([1/OR] – 1) × 100 when OR was >1.

[b] Conditional logistic models conditioned on matched pairs with additional adjustments.

[c] Model Adjustment

Age ≤17 years old group adjusted for days since last monovalent dose, number of outpatient and virtual visits, time since history of SARS-CoV-2 infection, history of SARS-CoV-2 molecular test, and medical center area.

Age 18-64 years old and age ≥65 years groups adjusted for days since last monovalent dose, number of emergency department visits, number of outpatient and virtual visits, time since history of SARS-CoV-2 infection, history of SARS-CoV-2 molecular test, BMI, Charlson comorbidity score, frailty index, medical center area, and number of monovalent doses prior to index date. Pregnancy added only to age 18-64 years old group.

[d] Medical center area dropped due to lack of model convergence.

[e] Time since history of SARS-CoV-2 infection dropped due to lack of model convergence.

[f] BMI dropped due to lack of model convergence.

[g] History of SARS-CoV-2 molecular test and Charlson comorbidity score dropped due to lack of model convergence.

Supplementary Table 6. Vaccine effectiveness of mRNA-1273 bivalent vaccine vs. unvaccinated against infection and severe outcomes with SARS-CoV-2 variants by age group

| | Test Positive | | Test Negative | | Odds Ratio (95% CI) | | VE (95% CI)[a] | |
|---|---|---|---|---|---|---|---|---|
| Subvariant/Age group/Outcome | mRNA-1273 bivalent vaccinated (%) | Unvaccinated (%) | mRNA-1273 bivalent vaccinated (%) | Unvaccinated (%) | Unadjusted | Adjusted[b,c] | Unadjusted | Adjusted[b,c] |
| BA.4/BA.5 | | | | | | | | |
| Aged ≤17 years | | | | | | | | |
| SARS-CoV-2 infection | 4 (0.5%) | 828 (99.5%) | 14 (0.6%) | 2482 (99.4%) | 0.857 (0.282, 2.604) | 0.826 (0.257, 2.656) | 14.3% (-61.6%, 71.8%) | 17.4% (-62.3%, 74.3%) |
| ED/UC encounters [d] | 2 (0.6%) | 326 (99.4%) | 2 (0.2%) | 982 (99.8%) | 3.000 (0.423, 21.30) | 5.974 (0.745, 47.90) | -66.7% (-95.3%, 57.7%) | -83.3% (-97.9%, 25.5%) |
| COVID-19 hospitalization | 0 (0.0%) | 6 (100.0%) | 1 (5.6%) | 17 (94.4%) | N/A | N/A | N/A | N/A |
| COVID-19 hospital death | 0 | 0 | 0 | 0 | N/A | N/A | N/A | N/A |
| Aged 18-64 years | | | | | | | | |
| SARS-CoV-2 infection [d] | 446 (22.7%) | 1518 (77.3%) | 1348 (22.9%) | 4544 (77.1%) | 0.988 (0.860, 1.134) | 0.993 (0.857, 1.150) | 1.2% (-11.8%, 14.0%) | 0.7% (-13.0%, 14.3%) |
| ED/UC encounters [d] | 100 (21.1%) | 375 (78.9%) | 353 (24.8%) | 1072 (75.2%) | 0.766 (0.578, 1.016) | 0.707 (0.523, 0.955) | 23.4% (-1.6%, 42.2%) | 29.3% (4.5%, 47.7%) |
| COVID-19 hospitalization [d,e,f] | 1 (5.6%) | 17 (94.4%) | 16 (29.6%) | 38 (70.4%) | 0.114 (0.013, 0.978) | 0.089 (0.009, 0.893) | 88.6% (2.2%, 98.7%) | 91.1% (10.7%, 99.1%) |
| COVID-19 hospital death | 0 | 0 | 0 | 0 | N/A | N/A | N/A | N/A |
| Aged ≥65 years | | | | | | | | |
| SARS-CoV-2 infection | 301 (50.1%) | 300 (49.9%) | 1116 (61.9%) | 687 (38.1%) | 0.557 (0.453, 0.684) | 0.581 (0.460, 0.733) | 44.3% (31.6%, 54.7%) | 41.9% (26.7%, 54.0%) |
| ED/UC encounters | 116 (36.3%) | 204 (63.8%) | 565 (58.9%) | 395 (41.1%) | 0.329 (0.245, 0.441) | 0.298 (0.208, 0.426) | 67.1% (55.9%, 75.5%) | 70.2% (57.4%, 79.2%) |
| COVID-19 hospitalization [d] | 23 (23.7%) | 74 (76.3%) | 156 (53.6%) | 135 (46.4%) | 0.224 (0.126, 0.400) | 0.260 (0.132, 0.509) | 77.6% (60.0%, 87.4%) | 74.0% (49.1%, 86.8%) |
| COVID-19 hospital death [d,e,g] | 2 (22.2%) | 7 (77.8%) | 10 (37.0%) | 17 (63.0%) | 0.518 (0.097, 2.755) | 0.263 (0.038, 1.825) | 48.2% (-63.7%, 90.3%) | 73.7% (-45.2%, 96.2%) |
| XBB | | | | | | | | |
| Aged ≤17 years | | | | | | | | |
| SARS-CoV-2 infection | 4 (1.2%) | 319 (98.8%) | 14 (1.4%) | 955 (98.6%) | 0.852 (0.275, 2.640) | 0.980 (0.296, 3.250) | 14.8% (-62.1%, 72.5%) | 2.0% (-69.2%, 70.4%) |
| ED/UC encounters | 0 (0.0%) | 183 (100.0%) | 10 (1.8%) | 539 (98.2%) | N/A | N/A | N/A | N/A |
| COVID-19 hospitalization | 0 (0.0%) | 2 (100.0%) | 0 (0.0%) | 6 (100.0%) | N/A | N/A | N/A | N/A |
| COVID-19 hospital death | 0 | 0 | 0 | 0 | N/A | N/A | N/A | N/A |
| Aged 18-64 years | | | | | | | | |
| SARS-CoV-2 infection [d] | 454 (49.6%) | 461 (50.4%) | 1084 (39.5%) | 1661 (60.5%) | 1.599 (1.360, 1.880) | 1.648 (1.384, 1.962) | -37.5% (-46.8%, -26.5%) | -39.3% (-49.0%, -27.8%) |

| | | | | | | | | |
|---|---|---|---|---|---|---|---|---|
| ED/UC encounters [d] | 150 (43.1%) | 198 (56.9%) | 399 (38.2%) | 645 (61.8%) | 1.268 (0.971, 1.657) | 1.357 (1.019, 1.806) | -21.2% (-39.6%, 2.9%) | -26.3% (-44.6%, -1.9%) |
| COVID-19 hospitalization [d,e,f] | 4 (26.7%) | 11 (73.3%) | 18 (40.0%) | 27 (60.0%) | 0.347 (0.060, 1.990) | 0.353 (0.043, 2.928) | 65.3% (-49.7%, 94.0%) | 64.7% (-65.8%, 95.7%) |
| COVID-19 hospital death | 1 (100.0%) | 0 (0.0%) | 3 (100.0%) | 0 (0.0%) | N/A | N/A | N/A | N/A |
| Aged ≥65 years | | | | | | | | |
| SARS-CoV-2 infection [d] | 341 (74.3%) | 118 (25.7%) | 1025 (74.4%) | 352 (25.6%) | 0.992 (0.779, 1.265) | 0.901 (0.695, 1.169) | 0.8% (-20.9%, 22.1%) | 9.9% (-14.5%, 30.5%) |
| ED/UC encounters [d] | 193 (68.0%) | 91 (32.0%) | 623 (73.1%) | 229 (26.9%) | 0.781 (0.584, 1.044) | 0.725 (0.530, 0.992) | 21.9% (-4.3%, 41.6%) | 27.5% (0.8%, 47.0%) |
| COVID-19 hospitalization [d,e] | 36 (47.4%) | 40 (52.6%) | 176 (77.2%) | 52 (22.8%) | 0.282 (0.163, 0.489) | 0.267 (0.148, 0.482) | 71.8% (51.1%, 83.7%) | 73.3% (51.8%, 85.2%) |
| COVID-19 hospital death | 3 (75.0%) | 1 (25.0%) | 10 (83.3%) | 2 (16.7%) | 0.577 (0.033, 10.25) | N/A | 42.3% (-90.2%, 96.7%) | N/A |

BMI= body mass index, ED/UC = emergency department/urgent care, OR = odds ratio, VE = vaccine effectiveness, N/A = not applicable

[a] Calculated as (1 – OR) x 100 when OR was ≤1, and ([1/OR] – 1) × 100 when OR was >1.

[b] Conditional logistic models conditioned on matched pairs with adjustments.

[c] Model Adjustment

Age ≤17 years old group adjusted for time since history of SARS-CoV-2 infection, history of SARS-CoV-2 molecular test, number of outpatient and virtual visits, specimen type and medical center area.

Age 18-64 years old and age ≥65 years groups adjusted for time since history of SARS-CoV-2 infection, history of SARS-CoV-2 molecular test, number of outpatient and virtual visits, BMI, specimen type and medical center area. Pregnancy added only to age 18-64 years old group.

[d] Medical center area dropped due to lack of model convergence.

[e] BMI and specimen type dropped due to lack of model convergence.

[f] Pregnancy dropped due to lack of model convergence.

[g] History of SARS-CoV-2 molecular test dropped due to lack of model convergence.

Supplementary Table 7. Vaccine effectiveness of mRNA-1273 bivalent vaccine vs. ≥2 monovalent mRNA vaccines against infection and severe outcomes with SARS-CoV-2 variants among immunocompromised patients

| Subvariant/Outcome | Test Positive | | Test Negative | | Odds Ratio (95% CI) | | rVE (95% CI)[a] | |
|---|---|---|---|---|---|---|---|---|
| | mRNA-1273 bivalent vaccinated (%) | ≥2 doses monovalent mRNA vaccinated (%) | mRNA-1273 bivalent vaccinated (%) | ≥2 doses monovalent mRNA vaccinated (%) | Unadjusted | Adjusted[b] | Unadjusted | Adjusted[b] |
| BA.4/BA.5 | | | | | | | | |
| SARS-CoV-2 infection | 42 (14.1%) | 255 (85.9%) | 227 (16.5%) | 1151 (83.5%) | 0.835 (0.585, 1.192) | 0.669 (0.439, 1.019) | 16.5% (-16.1%, 41.5%) | 33.1% (-1.9%, 56.1%) |
| ED/UC encounters [c,d] | 23 (14.6%) | 135 (85.4%) | 246 (16.2%) | 1271 (83.8%) | 0.880 (0.554, 1.398) | 0.879 (0.516, 1.496) | 12.0% (-28.5%, 44.6%) | 12.1% (-33.1%, 48.4%) |
| COVID-19 hospitalization [c,d] | 4 (9.8%) | 37 (90.2%) | 265 (16.2%) | 1369 (83.8%) | 0.558 (0.197, 1.580) | 0.375 (0.120, 1.169) | 44.2% (-36.7%, 80.3%) | 62.5% (-14.5%, 88.0%) |
| COVID-19 hospital death [c,d,e] | 1 (33.3%) | 2 (66.7%) | 268 (16.0%) | 1404 (84.0%) | 2.619 (0.237, 28.99) | 2.409 (0.197, 29.51) | -61.8% (-96.6%, 76.3%) | -58.5% (-96.6%, 80.3%) |
| XBB | | | | | | | | |
| SARS-CoV-2 infection [c] | 48 (27.3%) | 128 (72.7%) | 162 (20.5%) | 628 (79.5%) | 1.454 (1.000, 2.113) | 1.017 (0.658, 1.570) | -31.2% (-52.7%, 0.0%) | -1.6% (-36.3%, 34.2%) |
| ED/UC encounters [c] | 27 (25.5%) | 79 (74.5%) | 183 (21.3%) | 677 (78.7%) | 1.264 (0.793, 2.016) | 0.963 (0.559, 1.660) | -20.9% (-50.4%, 20.7%) | 3.7% (-39.7%, 44.1%) |
| COVID-19 hospitalization [c,d] | 4 (12.9%) | 27 (87.1%) | 206 (22.0%) | 729 (78.0%) | 0.524 (0.181, 1.515) | 0.327 (0.102, 1.043) | 47.6% (-34.0%, 81.9%) | 67.3% (-4.2%, 89.8%) |
| COVID-19 hospital death [c,d,e] | 1 (33.3%) | 2 (66.7%) | 209 (21.7%) | 754 (78.3%) | 1.804 (0.163, 19.99) | 1.246 (0.101, 15.44) | -44.6% (-95.0%, 83.7%) | -19.7% (-93.5%, 89.9%) |

BMI= body mass index, ED/UC = emergency department/urgent care, OR = odds ratio, rVE = relative vaccine effectiveness
[a] Calculated as (1 – OR) x 100 when OR was ≤1, and ([1/OR] – 1) × 100 when OR was >1.
[b] Unconditional logistic regression models adjusted for age, sex, race/ethnicity, month of specimen collection, days since last monovalent dose, time since history of SARS-CoV-2 infection, history of SARS-CoV-2 molecular test, number of outpatient and virtual visits, number of emergency department visits, BMI, Charlson comorbidity score, frailty index, pregnancy, medical center area, and number of monovalent doses prior to index date.
[c] Medical center area dropped due to lack of model convergence.
[d] BMI and pregnancy dropped due to lack of model convergence.
[e] Month of specimen collection, Charlson comorbidity score, frailty index, time since history of SARS-CoV-2 infection, history of SARS-CoV-2 molecular test, number of outpatient and virtual visits, number of emergency department visits, and number of monovalent doses prior to index date dropped due to lack of model convergence.

Supplementary Table 8. Vaccine effectiveness of mRNA-1273 bivalent vaccine vs. unvaccinated against infection and severe outcomes with SARS-CoV-2 variants among immunocompromised patients

| | Test Positive | | Test Negative | | Odds Ratio (95% CI) | | VE (95% CI)[b] | |
|---|---|---|---|---|---|---|---|---|
| Subvariant/Outcome | mRNA-1273 bivalent vaccinated (%) | Unvaccinated (%) | mRNA-1273 bivalent vaccinated (%) | Unvaccinated (%) | Unadjusted | Adjusted[a] | Unadjusted | Adjusted[c] |
| BA.4/BA.5 | | | | | | | | |
| SARS-CoV-2 infection | 42 (36.5%) | 73 (63.5%) | 179 (39.9%) | 270 (60.1%) | 0.868 (0.568, 1.326) | 0.530 (0.297, 0.949) | 13.2% (-24.6%, 43.2%) | 47.0% (5.1%, 70.3%) |
| ED/UC encounters | 23 (36.5%) | 40 (63.5%) | 198 (39.5%) | 303 (60.5%) | 0.880 (0.511, 1.515) | 0.614 (0.314, 1.202) | 12.0% (-34.0%, 48.9%) | 38.6% (-16.8%, 68.6%) |
| COVID-19 hospitalization [c,d,e] | 4 (26.7%) | 11 (73.3%) | 217 (39.5%) | 332 (60.5%) | 0.556 (0.175, 1.770) | 0.225 (0.063, 0.798) | 44.4% (-43.5%, 82.5%) | 77.5% (20.2%, 93.7%) |
| COVID-19 hospital death | 1 (100.0%) | 0 (0.0%) | 220 (39.1%) | 343 (60.9%) | N/A | N/A | N/A | N/A |
| XBB | | | | | | | | |
| SARS-CoV-2 infection[c] | 48 (60.8%) | 31 (39.2%) | 171 (53.3%) | 150 (46.7%) | 1.358 (0.822, 2.244) | 1.178 (0.618, 2.246) | -26.4% (-55.4%, 17.8%) | -15.1% (-55.5%, 38.2%) |
| ED/UC encounters [c] | 27 (58.7%) | 19 (41.3%) | 192 (54.2%) | 162 (45.8%) | 1.199 (0.643, 2.236) | 0.993 (0.459, 2.145) | -16.6% (-55.3%, 35.7%) | 0.7% (-53.4%, 54.1%) |
| COVID-19 hospitalization[c,d] | 4 (33.3%) | 8 (66.7%) | 215 (55.4%) | 173 (44.6%) | 0.402 (0.119, 1.358) | 0.306 (0.080, 1.177) | 59.8% (-26.4%, 88.1%) | 69.4% (-15.1%, 92.0%) |
| COVID-19 hospital death | 1 (100.0%) | 0 (0.0%) | 218 (54.6%) | 181 (45.4%) | N/A | N/A | N/A | N/A |

BMI= body mass index, ED/UC = emergency department/urgent care, OR = odds ratio, VE = vaccine effectiveness, N/A = not applicable
[a] Calculated as (1 – OR) x 100 when OR was ≤1, and ([1/OR] – 1) × 100 when OR was >1.
[b] Unconditional logistic regression models adjusted for age, sex, race/ethnicity, month of specimen collection, time since history of SARS-CoV-2 infection, history of SARS-CoV-2 molecular test, number of outpatient and virtual visits, BMI, pregnancy, and specimen type.
[c] Pregnancy dropped due to lack of model convergence.
[d] BMI and specimen type dropped due to lack of model convergence.
[e] Month of specimen collection and history of SARS-CoV-2 molecular test dropped due to lack of model convergence.

Supplementary Table 9. Vaccine effectiveness of mRNA-1273 bivalent vaccine vs. ≥2 monovalent mRNA vaccines against infection and severe outcomes with SARS-CoV-2 variants among those with history of SARS-CoV-2 infection

| | Test Positive | | Test Negative | | Odds Ratio (95% CI) | | rVE (95% CI)[a] | |
|---|---|---|---|---|---|---|---|---|
| **Subvariant/Outcome** | **mRNA-1273 bivalent vaccinated (%)** | **≥2 doses monovalent mRNA vaccinated (%)** | **mRNA-1273 bivalent vaccinated (%)** | **≥2 doses monovalent mRNA vaccinated (%)** | **Unadjusted** | **Adjusted[b]** | **Unadjusted** | **Adjusted[b]** |
| BA.4/BA.5 | | | | | | | | |
| SARS-CoV-2 infection | 91 (5.9%) | 1453 (94.1%) | 1014 (12.7%) | 6942 (87.3%) | 0.429 (0.343, 0.535) | 0.423 (0.333, 0.537) | 57.1% (46.5%, 65.7%) | 57.7% (46.3%, 66.7%) |
| ED/UC encounters[c] | 23 (6.9%) | 308 (93.1%) | 1082 (11.8%) | 8087 (88.2%) | 0.558 (0.364, 0.857) | 0.456 (0.289, 0.720) | 44.2% (14.3%, 63.6%) | 54.4% (28.0%, 71.1%) |
| COVID-19 hospitalization [c,d] | 1 (3.8%) | 25 (96.2%) | 1104 (11.7%) | 8370 (88.3%) | 0.303 (0.041, 2.240) | 0.154 (0.020, 1.207) | 69.7% (-55.4%, 95.9%) | 84.6% (-17.1%, 98.0%) |
| COVID-19 hospital death | 0 (0.0%) | 2 (100.0%) | 1105 (11.6%) | 8393 (88.4%) | N/A | N/A | N/A | N/A |
| XBB | | | | | | | | |
| SARS-CoV-2 infection | 196 (15.5%) | 1071 (84.5%) | 653 (15.6%) | 3536 (84.4%) | 0.991 (0.833, 1.179) | 0.964 (0.795, 1.169) | 0.9% (-15.2%, 16.7%) | 3.6% (-14.4%, 20.5%) |
| ED/UC encounters[c] | 72 (16.1%) | 376 (83.9%) | 777 (15.5%) | 4231 (84.5%) | 1.043 (0.801, 1.357) | 0.845 (0.632, 1.128) | -4.1% (-26.3%, 19.9%) | 15.5% (-11.4%, 36.8%) |
| COVID-19 hospitalization [c,d,e] | 5 (12.8%) | 34 (87.2%) | 844 (15.6%) | 4573 (84.4%) | 0.797 (0.311, 2.043) | 0.382 (0.140, 1.042) | 20.3% (-51.1%, 68.9%) | 61.8% (-4.0%, 86.0%) |
| COVID-19 hospital death | 0 (0.0%) | 1 (100.0%) | 849 (15.6%) | 4606 (84.4%) | N/A | N/A | N/A | N/A |

BMI= body mass index, ED/UC = emergency department/urgent care, OR = odds ratio, rVE = relative vaccine effectiveness, N/A = not applicable
[a] Calculated as (1 – OR) x 100 when OR was ≤1, and ([1/OR] – 1) × 100 when OR was >1.
[b] Unconditional logistic regression models adjusted for age, sex, race/ethnicity, month of specimen collection, days since last monovalent dose, time since history of SARS-CoV-2 infection, history of SARS-CoV-2 molecular test, number of outpatient and virtual visits, number of emergency department visits, BMI, Charlson comorbidity score, frailty index, pregnancy, medical center area, and number of monovalent doses prior to index date.
[c] Medical center area dropped due to lack of model convergence.
[d] BMI and pregnancy dropped due to lack of model convergence.
[e] Frailty index dropped due to lack of model convergence.

Supplementary Table 10. Vaccine effectiveness of mRNA-1273 bivalent vaccine vs. unvaccinated against infection and severe outcomes with SARS-CoV-2 variants among those with history of SARS-CoV-2 infection

| | Test Positive | | Test Negative | | Odds Ratio (95% CI) | | VE (95% CI)[a] | |
|---|---|---|---|---|---|---|---|---|
| Subvariant/Outcome | mRNA-1273 bivalent vaccinated (%) | Unvaccinated (%) | mRNA-1273 bivalent vaccinated (%) | Unvaccinated (%) | Unadjusted | Adjusted[b] | Unadjusted | Adjusted[b] |
| BA.4/BA.5 | | | | | | | | |
| SARS-CoV-2 infection | 91 (9.3%) | 886 (90.7%) | 735 (19.5%) | 3043 (80.5%) | 0.425 (0.338, 0.535) | 0.482 (0.372, 0.625) | 57.5% (46.5%, 66.2%) | 51.8% (37.5%, 62.8%) |
| ED/UC encounters | 23 (10.6%) | 195 (89.4%) | 803 (17.7%) | 3734 (82.3%) | 0.548 (0.354, 0.851) | 0.402 (0.246, 0.656) | 45.2% (14.9%, 64.6%) | 59.8% (34.4%, 75.4%) |
| COVID-19 hospitalization [c,d] | 1 (9.1%) | 10 (90.9%) | 825 (17.4%) | 3919 (82.6%) | 0.475 (0.061, 3.716) | 0.140 (0.016, 1.195) | 52.5% (-73.1%, 93.9%) | 86.0% (-16.3%, 98.4%) |
| COVID-19 hospital death | 0 (0.0%) | 1 (100.0%) | 826 (17.4%) | 3928 (82.6%) | N/A | N/A | N/A | N/A |
| XBB | | | | | | | | |
| SARS-CoV-2 infection | 196 (38.7%) | 311 (61.3%) | 632 (37.2%) | 1066 (62.8%) | 1.063 (0.867, 1.304) | 1.253 (0.972, 1.617) | -5.9% (-23.3%, 13.3%) | -20.2% (-38.1%, 2.8%) |
| ED/UC encounters[c] | 72 (36.5%) | 125 (63.5%) | 756 (37.6%) | 1252 (62.4%) | 0.954 (0.704, 1.292) | 0.787 (0.550, 1.128) | 4.6% (-22.6%, 29.6%) | 21.3% (-11.3%, 45.0%) |
| COVID-19 hospitalization [c,d,e] | 5 (41.7%) | 7 (58.3%) | 823 (37.5%) | 1370 (62.5%) | 1.189 (0.376, 3.759) | 0.342 (0.095, 1.233) | -15.9% (-73.4%, 62.4%) | 65.8% (-18.9%, 90.5%) |
| COVID-19 hospital death | 0 | 0 | 828 (37.6%) | 1377 (62.4%) | N/A | N/A | N/A | N/A |

BMI= body mass index, ED/UC = emergency department/urgent care, OR = odds ratio, VE = vaccine effectiveness, N/A = not applicable

[a] Calculated as (1 – OR) x 100 when OR was ≤1, and ([1/OR] – 1) × 100 when OR was >1.

[b] Unconditional logistic regression models adjusted for age, sex, race/ethnicity, month of specimen collection, time since history of SARS-CoV-2 infection, history of SARS-CoV-2 molecular test, number of outpatient and virtual visits, BMI, pregnancy, specimen type, and medical center area.

[c] Medical center area dropped due to lack of model convergence.

[d] BMI and specimen type dropped due to lack of model convergence.

[e] History of SARS-CoV-2 molecular test, number of outpatient and virtual visits, and pregnancy dropped due to lack of model convergence.

Supplementary Table 11. Vaccine effectiveness of mRNA-1273 bivalent vaccine vs. ≥2 monovalent mRNA vaccines against infection and severe outcomes with SARS-CoV-2 variants by time since vaccination, using SGTF data to assign unidentified subvariants

| Subvariant/Time since vaccination | Test Positive | | Test Negative | | Odds Ratio (95% CI) | | rVE (95% CI)[a] | |
|---|---|---|---|---|---|---|---|---|
| | mRNA-1273 bivalent vaccinated (%) | ≥2 doses monovalent mRNA vaccinated (%) | mRNA-1273 bivalent vaccinated (%) | ≥2 doses monovalent mRNA vaccinated (%) | Unadjusted | Adjusted[b,c] | Unadjusted | Adjusted[b,c] |
| BA.4/BA.5 | | | | | | | | |
| SARS-CoV-2 infection | 1375 (11.1%) | 10957 (88.9%) | 5678 (15.3%) | 31318 (84.7%) | 0.661 (0.618, 0.706) | 0.590 (0.549, 0.634) | 33.9% (29.4%, 38.2%) | 41.0% (36.6%, 45.1%) |
| 14-60 days | 788 (6.7%) | 10957 (93.3%) | 3624 (10.4%) | 31318 (89.6%) | 0.621 (0.574, 0.673) | 0.527 (0.483, 0.574) | 37.9% (32.7%, 42.6%) | 47.3% (42.6%, 51.7%) |
| 61-120 days | 535 (4.7%) | 10957 (95.3%) | 1878 (5.7%) | 31318 (94.3%) | 0.814 (0.738, 0.899) | 0.665 (0.598, 0.741) | 18.6% (10.1%, 26.2%) | 33.5% (25.9%, 40.2%) |
| 121-180 days | 51 (0.5%) | 10957 (99.5%) | 175 (0.6%) | 31318 (99.4%) | 0.833 (0.609, 1.139) | 0.721 (0.519, 1.001) | 16.7% (-12.2%, 39.1%) | 27.9% (-0.1%, 48.1%) |
| >180 days | 1 (0.0%) | 10957 (100.0%) | 1 (0.0%) | 31318 (100.0%) | 2.858 (0.179, 45.70) | 1.863 (0.109, 31.74) | -65.0% (-97.8%, 82.1%) | -46.3% (-96.8%, 89.1%) |
| ED/UC encounters | 364 (11.9%) | 2700 (88.1%) | 1599 (17.4%) | 7593 (82.6%) | 0.602 (0.529, 0.686) | 0.498 (0.429, 0.577) | 39.8% (31.4%, 47.1%) | 50.2% (42.3%, 57.1%) |
| 14-60 days [d] | 210 (7.2%) | 2700 (92.8%) | 984 (11.5%) | 7593 (88.5%) | 0.600 (0.514, 0.701) | 0.525 (0.444, 0.621) | 40.0% (29.9%, 48.6%) | 47.5% (37.9%, 55.6%) |
| 61-120 days [d] | 136 (4.8%) | 2700 (95.2%) | 551 (6.8%) | 7593 (93.2%) | 0.694 (0.572, 0.842) | 0.609 (0.494, 0.751) | 30.6% (15.8%, 42.8%) | 39.1% (24.9%, 50.6%) |
| 121-180 days [d] | 17 (0.6%) | 2700 (99.4%) | 64 (0.8%) | 7593 (99.2%) | 0.747 (0.437, 1.277) | 0.699 (0.398, 1.226) | 25.3% (-21.7%, 56.3%) | 30.1% (-18.4%, 60.2%) |
| >180 days [d] | 1 (0.0%) | 2700 (100.0%) | 0 (0.0%) | 7593 (100.0%) | N/A | N/A | N/A | N/A |
| COVID-19 hospitalization[d] | 28 (10.4%) | 241 (89.6%) | 208 (25.8%) | 599 (74.2%) | 0.307 (0.198, 0.478) | 0.372 (0.222, 0.624) | 69.3% (52.2%, 80.2%) | 62.8% (37.6%, 77.8%) |
| 14-60 days [d] | 13 (5.1%) | 241 (94.9%) | 133 (18.2%) | 599 (81.8%) | 0.243 (0.135, 0.438) | 0.318 (0.170, 0.597) | 75.7% (56.2%, 86.5%) | 68.2% (40.3%, 83.0%) |
| 61-120 days [d] | 14 (5.5%) | 241 (94.5%) | 70 (10.5%) | 599 (89.5%) | 0.497 (0.275, 0.899) | 0.593 (0.301, 1.170) | 50.3% (10.1%, 72.5%) | 40.7% (-14.5%, 69.9%) |
| 121-180 days [d] | 0 (0.0%) | 241 (100.0%) | 5 (0.8%) | 599 (99.2%) | N/A | N/A | N/A | N/A |
| >180 days [d] | 1 (0.4%) | 241 (99.6%) | 0 (0.0%) | 599 (100.0%) | N/A | N/A | N/A | N/A |
| COVID-19 hospital death [d,e] | 2 (11.8%) | 15 (88.2%) | 13 (25.5%) | 38 (74.5%) | 0.358 (0.069, 1.869) | 0.469 (0.035, 6.337) | 64.2% (-46.5%, 93.1%) | 53.1% (-84.2%, 96.5%) |
| XBB | | | | | | | | |
| SARS-CoV-2 infection | 1166 (21.2%) | 4334 (78.8%) | 3132 (19.0%) | 13368 (81.0%) | 1.160 (1.073, 1.255) | 0.972 (0.891, 1.060) | -13.8% (-20.3%, -6.8%) | 2.8% (-5.7%, 10.9%) |
| 14-60 days | 106 (2.4%) | 4334 (97.6%) | 442 (3.2%) | 13368 (96.8%) | 0.740 (0.597, 0.917) | 0.587 (0.469, 0.736) | 26.0% (8.3%, 40.3%) | 41.3% (26.4%, 53.1%) |
| 61-120 days | 303 (6.5%) | 4334 (93.5%) | 984 (6.9%) | 13368 (93.1%) | 0.950 (0.831, 1.085) | 0.771 (0.667, 0.890) | 5.0% (-7.8%, 16.9%) | 22.9% (11.0%, 33.3%) |
| 121-180 days | 401 (8.5%) | 4334 (91.5%) | 970 (6.8%) | 13368 (93.2%) | 1.275 (1.129, 1.440) | 1.098 (0.963, 1.252) | -21.6% (-30.5%, -11.5%) | -9.0% (-20.1%, 3.7%) |
| >180 days | 356 (7.6%) | 4334 (92.4%) | 736 (5.2%) | 13368 (94.8%) | 1.492 (1.309, 1.701) | 1.273 (1.101, 1.472) | -33.0% (-41.2%, -23.6%) | -21.5% (-32.1%, -9.2%) |
| ED/UC encounters | 408 (21.2%) | 1513 (78.8%) | 1255 (21.8%) | 4508 (78.2%) | 0.967 (0.849, 1.101) | 0.764 (0.660, 0.886) | 3.3% (-9.2%, 15.1%) | 23.6% (11.4%, 34.0%) |

| | | | | | | | | |
|---|---|---|---|---|---|---|---|---|
| 14-60 days [d] | 24 (1.6%) | 1513 (98.4%) | 176 (3.8%) | 4508 (96.2%) | 0.406 (0.264, 0.625) | 0.310 (0.199, 0.484) | 59.4% (37.5%, 73.6%) | 69.0% (51.6%, 80.1%) |
| 61-120 days [d] | 107 (6.6%) | 1513 (93.4%) | 342 (7.1%) | 4508 (92.9%) | 0.932 (0.745, 1.167) | 0.730 (0.573, 0.929) | 6.8% (-14.3%, 25.5%) | 27.0% (7.1%, 42.7%) |
| 121-180 days [d] | 146 (8.8%) | 1513 (91.2%) | 404 (8.2%) | 4508 (91.8%) | 1.077 (0.883, 1.313) | 0.931 (0.753, 1.150) | -7.1% (-23.8%, 11.7%) | 6.9% (-13.1%, 24.7%) |
| >180 days [d] | 131 (8.0%) | 1513 (92.0%) | 333 (6.9%) | 4508 (93.1%) | 1.172 (0.950, 1.447) | 0.983 (0.780, 1.240) | -14.7% (-30.9%, 5.0%) | 1.7% (-19.3%, 22.0%) |
| COVID-19 hospitalization [d] | 40 (18.9%) | 172 (81.1%) | 209 (32.9%) | 427 (67.1%) | 0.482 (0.329, 0.705) | 0.401 (0.255, 0.630) | 51.8% (29.5%, 67.1%) | 59.9% (37.0%, 74.5%) |
| 14-60 days [d] | 2 (1.1%) | 172 (98.9%) | 27 (5.9%) | 427 (94.1%) | 0.184 (0.043, 0.782) | 0.128 (0.027, 0.596) | 81.6% (21.8%, 95.7%) | 87.2% (40.4%, 97.3%) |
| 61-120 days [d] | 7 (3.9%) | 172 (96.1%) | 54 (11.2%) | 427 (88.8%) | 0.322 (0.144, 0.721) | 0.236 (0.098, 0.568) | 67.8% (27.9%, 85.6%) | 76.4% (43.2%, 90.2%) |
| 121-180 days [d] | 14 (7.5%) | 172 (92.5%) | 72 (14.4%) | 427 (85.6%) | 0.483 (0.265, 0.879) | 0.433 (0.224, 0.836) | 51.7% (12.1%, 73.5%) | 56.7% (16.4%, 77.6%) |
| >180 days [d] | 17 (9.0%) | 172 (91.0%) | 56 (11.6%) | 427 (88.4%) | 0.754 (0.426, 1.334) | 0.643 (0.336, 1.232) | 24.6% (-25.0%, 57.4%) | 35.7% (-18.8%, 66.4%) |
| COVID-19 hospital death [d,e,f] | 4 (28.6%) | 10 (71.4%) | 17 (40.5%) | 25 (59.5%) | 0.602 (0.166, 2.174) | 0.682 (0.076, 6.133) | 39.8% (-54.0%, 83.4%) | 31.8% (-83.7%, 92.4%) |

BMI= body mass index, ED/UC = emergency department/urgent care, OR = odds ratio, rVE = relative vaccine effectiveness, SGTF = S gene target failure, N/A = not applicable

[a] Calculated as (1 – OR) x 100 when OR was ≤1, and ([1/OR] – 1) × 100 when OR was >1

[b] Applied conditional logistic models conditioned on matched pairs for SARS-CoV-2 infection, ED/UC encounters, hospitalization and hospital death outcomes. Models for time since vaccination analyses are unconditional logistic models, adjusted for age, sex, race/ethnicity, and month of specimen collection, in addition to the covariates listed below.

[c] Adjusted for days since last monovalent dose, time since history of SARS-CoV-2 infection, history of SARS-CoV-2 molecular test, number of outpatient and virtual visits, number of emergency department visits, BMI, Charlson comorbidity score, frailty index, pregnancy, medical center area, and number of monovalent doses prior to index date.

[d] Medical center area dropped due to lack of model convergence.

[e] BMI, Charlson comorbidity score, frailty index, and pregnancy dropped due to lack of model convergence.

[f] Number of monovalent doses prior to index date dropped due to lack of model convergence.

Supplementary Table 12. Vaccine effectiveness of mRNA-1273 bivalent vaccine vs. unvaccinated against infection and severe outcomes with SARS-CoV-2 variants by time since vaccination, using SGTF data to assign unidentified subvariants

| | Test Positive | | Test Negative | | Odds Ratio (95% CI) | | VE (95% CI)[a] | |
|---|---|---|---|---|---|---|---|---|
| **Subvariant/Time since vaccination** | **mRNA-1273 bivalent vaccinated (%)** | **Unvaccinated (%)** | **mRNA-1273 bivalent vaccinated (%)** | **Unvaccinated (%)** | **Unadjusted** | **Adjusted [b,c]** | **Unadjusted** | **Adjusted [b,c]** |
| BA.4/BA.5 | | | | | | | | |
| SARS-CoV-2 infection[d] | 1376 (22.4%) | 4769 (77.6%) | 4430 (24.0%) | 14005 (76.0%) | 0.867 (0.795, 0.944) | 0.852 (0.778, 0.933) | 13.3% (5.6%, 20.5%) | 14.8% (6.7%, 22.2%) |
| 14-60 days [d] | 789 (14.2%) | 4769 (85.8%) | 2811 (16.7%) | 14005 (83.3%) | 0.824 (0.757, 0.898) | 0.761 (0.689, 0.841) | 17.6% (10.2%, 24.3%) | 23.9% (15.9%, 31.1%) |
| 61-120 days [d] | 535 (10.1%) | 4769 (89.9%) | 1456 (9.4%) | 14005 (90.6%) | 1.079 (0.972, 1.198) | 0.981 (0.870, 1.107) | -7.3% (-16.5%, 2.8%) | 1.9% (-9.6%, 13.0%) |
| 121-180 days [d] | 51 (1.1%) | 4769 (98.9%) | 161 (1.1%) | 14005 (98.9%) | 0.930 (0.678, 1.277) | 0.969 (0.687, 1.366) | 7.0% (-21.7%, 32.2%) | 3.1% (-26.8%, 31.3%) |
| >180 days [d] | 1 (0.0%) | 4769 (100.0%) | 2 (0.0%) | 14005 (100.0%) | 1.469 (0.133, 16.20) | 1.339 (0.083, 21.51) | -31.9% (-93.8%, 86.7%) | -25.3% (-95.4%, 91.7%) |
| ED/UC encounters | 365 (24.1%) | 1152 (75.9%) | 1260 (27.7%) | 3291 (72.3%) | 0.748 (0.634, 0.883) | 0.528 (0.438, 0.637) | 25.2% (11.7%, 36.6%) | 47.2% (36.3%, 56.2%) |
| 14-60 days | 211 (15.5%) | 1152 (84.5%) | 784 (19.2%) | 3291 (80.8%) | 0.769 (0.651, 0.908) | 0.505 (0.412, 0.620) | 23.1% (9.2%, 34.9%) | 49.5% (38.0%, 58.8%) |
| 61-120 days | 136 (10.6%) | 1152 (89.4%) | 426 (11.5%) | 3291 (88.5%) | 0.912 (0.743, 1.119) | 0.604 (0.473, 0.771) | 8.8% (-10.6%, 25.7%) | 39.6% (22.9%, 52.7%) |
| 121-180 days | 17 (1.5%) | 1152 (98.5%) | 48 (1.4%) | 3291 (98.6%) | 1.012 (0.580, 1.766) | 0.755 (0.399, 1.430) | -1.2% (-43.4%, 42.0%) | 24.5% (-30.1%, 60.1%) |
| >180 days | 1 (0.1%) | 1152 (99.9%) | 2 (0.1%) | 3291 (99.9%) | 1.429 (0.129, 15.77) | 0.737 (0.039, 14.10) | -30.0% (-93.7%, 87.1%) | 26.3% (-92.9%, 96.1%) |
| COVID-19 hospitalization[d] | 28 (21.4%) | 103 (78.6%) | 189 (48.1%) | 204 (51.9%) | 0.212 (0.123, 0.364) | 0.209 (0.110, 0.398) | 78.8% (63.6%, 87.7%) | 79.1% (60.2%, 89.0%) |
| 14-60 days [d] | 13 (11.2%) | 103 (88.8%) | 109 (34.8%) | 204 (65.2%) | 0.236 (0.127, 0.440) | 0.234 (0.118, 0.466) | 76.4% (56.0%, 87.3%) | 76.6% (53.4%, 88.2%) |
| 61-120 days [d] | 14 (12.0%) | 103 (88.0%) | 74 (26.6%) | 204 (73.4%) | 0.375 (0.202, 0.695) | 0.372 (0.187, 0.738) | 62.5% (30.5%, 79.8%) | 62.8% (26.2%, 81.3%) |
| 121-180 days [d] | 0 (0.0%) | 103 (100.0%) | 6 (2.9%) | 204 (97.1%) | N/A | N/A | N/A | N/A |
| >180 days [d] | 1 (1.0%) | 103 (99.0%) | 0 (0.0%) | 204 (100.0%) | N/A | N/A | N/A | N/A |
| COVID-19 hospital death [d,e,f] | 2 (20.0%) | 8 (80.0%) | 12 (40.0%) | 18 (60.0%) | 0.414 (0.082, 2.102) | 0.409 (0.077, 2.168) | 58.6% (-52.4%, 91.8%) | 59.1% (-53.9%, 92.3%) |
| XBB | | | | | | | | |
| SARS-CoV-2 infection[d] | 1166 (48.3%) | 1250 (51.7%) | 2995 (41.3%) | 4253 (58.7%) | 1.516 (1.353, 1.699) | 1.420 (1.258, 1.603) | -34.0% (-41.1%, -26.1%) | -29.6% (-37.6%, -20.5%) |
| 14-60 days [d] | 106 (7.8%) | 1250 (92.2%) | 414 (8.9%) | 4253 (91.1%) | 0.871 (0.697, 1.088) | 0.837 (0.658, 1.066) | 12.9% (-8.1%, 30.3%) | 16.3% (-6.2%, 34.2%) |
| 61-120 days [d] | 303 (19.5%) | 1250 (80.5%) | 899 (17.4%) | 4253 (82.6%) | 1.147 (0.992, 1.325) | 1.186 (1.004, 1.400) | -12.8% (-24.5%, 0.8%) | -15.7% (-28.6%, -0.4%) |
| 121-180 days [d] | 401 (24.3%) | 1250 (75.7%) | 948 (18.2%) | 4253 (81.8%) | 1.439 (1.260, 1.643) | 1.588 (1.358, 1.858) | -30.5% (-39.2%, -20.7%) | -37.0% (-46.2%, -26.4%) |

| | | | | | | | | |
|---|---|---|---|---|---|---|---|---|
| >180 days [d] | 356 (22.2%) | 1250 (77.8%) | 734 (14.7%) | 4253 (85.3%) | 1.651 (1.433, 1.901) | 1.990 (1.666, 2.376) | -39.4% (-47.4%, -30.2%) | -49.7% (-57.9%, -40.0%) |
| ED/UC encounters | 408 (44.4%) | 511 (55.6%) | 1172 (42.5%) | 1585 (57.5%) | 1.123 (0.933, 1.352) | 1.005 (0.821, 1.230) | -11.0% (-26.1%, 6.7%) | -0.5% (-18.7%, 17.9%) |
| 14-60 days | 24 (4.5%) | 511 (95.5%) | 154 (8.9%) | 1585 (91.1%) | 0.483 (0.311, 0.752) | 0.454 (0.284, 0.726) | 51.7% (24.8%, 68.9%) | 54.6% (27.4%, 71.6%) |
| 61-120 days | 107 (17.3%) | 511 (82.7%) | 326 (17.1%) | 1585 (82.9%) | 1.018 (0.801, 1.294) | 0.925 (0.705, 1.214) | -1.8% (-22.7%, 19.9%) | 7.5% (-17.6%, 29.5%) |
| 121-180 days | 146 (22.2%) | 511 (77.8%) | 377 (19.2%) | 1585 (80.8%) | 1.201 (0.968, 1.490) | 1.146 (0.886, 1.482) | -16.8% (-32.9%, 3.2%) | -12.7% (-32.5%, 11.4%) |
| >180 days | 131 (20.4%) | 511 (79.6%) | 315 (16.6%) | 1585 (83.4%) | 1.290 (1.028, 1.618) | 1.219 (0.927, 1.603) | -22.5% (-38.2%, -2.7%) | -18.0% (-37.6%, 7.3%) |
| COVID-19 hospitalization[d,e] | 40 (42.1%) | 55 (57.9%) | 197 (69.1%) | 88 (30.9%) | 0.281 (0.167, 0.474) | 0.259 (0.142, 0.471) | 71.9% (52.6%, 83.3%) | 74.1% (52.9%, 85.8%) |
| 14-60 days [d,e] | 2 (3.5%) | 55 (96.5%) | 37 (29.6%) | 88 (70.4%) | 0.086 (0.020, 0.373) | 0.063 (0.013, 0.295) | 91.4% (62.7%, 98.0%) | 93.7% (70.5%, 98.7%) |
| 61-120 days [d,e] | 7 (11.3%) | 55 (88.7%) | 58 (39.7%) | 88 (60.3%) | 0.193 (0.082, 0.453) | 0.143 (0.055, 0.373) | 80.7% (54.7%, 91.8%) | 85.7% (62.7%, 94.5%) |
| 121-180 days [d,e] | 14 (20.3%) | 55 (79.7%) | 55 (38.5%) | 88 (61.5%) | 0.407 (0.207, 0.801) | 0.364 (0.167, 0.796) | 59.3% (19.9%, 79.3%) | 63.6% (20.4%, 83.3%) |
| >180 days [d,e] | 17 (23.6%) | 55 (76.4%) | 47 (34.8%) | 88 (65.2%) | 0.579 (0.302, 1.107) | 0.562 (0.261, 1.208) | 42.1% (-9.7%, 69.8%) | 43.8% (-17.2%, 73.9%) |
| COVID-19 hospital death[d,e,f] | 4 (80.0%) | 1 (20.0%) | 13 (86.7%) | 2 (13.3%) | 0.577 (0.033, 10.25) | 0.690 (0.035, 13.55) | 42.3% (-90.2%, 96.7%) | 31.0% (-92.6%, 96.5%) |

BMI= body mass index, ED/UC = emergency department/urgent care, OR = odds ratio, VE = vaccine effectiveness, SGTF = S gene target failure, N/A = not applicable

[a] Calculated as (1 – OR) x 100 when OR was ≤1, and ([1/OR] – 1) × 100 when OR was >1.

[b] Applied conditional logistic models conditioned on matched pairs for SARS-CoV-2 infection, ED/UC encounters, hospitalization and hospital death outcomes. Models for time since vaccination analyses are unconditional logistic models, adjusted for age, sex, race/ethnicity, and month of specimen collection, in addition to the covariates listed below.

[c] Adjusted for time since history of SARS-CoV-2 infection, history of SARS-CoV-2 molecular test, number of outpatient and virtual visits, BMI, pregnancy, specimen type, and medical center area.

[d] Medical center area dropped due to lack of model convergence.

[e] Specimen type and pregnancy dropped due to lack of model convergence.

[f] BMI and time since history of SARS-CoV-2 infection dropped due to lack of model convergence.

Supplementary Table 13. Vaccine effectiveness of mRNA-1273 bivalent vaccine vs. ≥2 monovalent mRNA vaccines against infection and severe outcomes with XBB.1.5 subvariant

| | Test Positive | | Test Negative | | Odds Ratio (95% CI) | | rVE (95% CI)[a] | |
|---|---|---|---|---|---|---|---|---|
| **Outcome** | **mRNA-1273 bivalent vaccinated (%)** | **≥2 doses monovalent mRNA vaccinated (%)** | **mRNA-1273 bivalent vaccinated (%)** | **≥2 doses monovalent mRNA vaccinated (%)** | **Unadjusted** | **Adjusted[b]** | **Unadjusted** | **Adjusted[b]** |
| SARS-CoV-2 infection | 533 (20.7%) | 2045 (79.3%) | 1575 (20.4%) | 6159 (79.6%) | 1.021 (0.910, 1.144) | 0.827 (0.728, 0.939) | -2.0% (-12.6%, 9.0%) | 17.3% (6.1%, 27.2%) |
| ED/UC encounters | 230 (20.4%) | 898 (79.6%) | 784 (23.2%) | 2600 (76.8%) | 0.840 (0.708, 0.996) | 0.692 (0.571, 0.839) | 16.0% (0.4%, 29.2%) | 30.8% (16.1%, 42.9%) |
| COVID-19 hospitalization[c] | 25 (16.8%) | 124 (83.2%) | 143 (32.0%) | 304 (68.0%) | 0.432 (0.269, 0.695) | 0.375 (0.211, 0.667) | 56.8% (30.5%, 73.1%) | 62.5% (33.3%, 78.9%) |
| COVID-19 hospital death [c,d] | 1 (16.7%) | 5 (83.3%) | 6 (33.3%) | 12 (66.7%) | 0.361 (0.031, 4.214) | 0.199 (0.007, 5.664) | 63.9% (-76.3%, 96.9%) | 80.1% (-82.3%, 99.3%) |

BMI= body mass index, ED/UC = emergency department/urgent care, OR = odds ratio, rVE = relative vaccine effectiveness

[a] Calculated as (1 – OR) x 100 when OR was ≤1, and ([1/OR] – 1) × 100 when OR was >1.

[b] Applied conditional logistic models conditioned on matched pairs, additionally adjusted for days since last monovalent dose, time since history of SARS-CoV-2 infection, history of SARS-CoV-2 molecular test, number of outpatient and virtual visits, number of emergency department visits, BMI, Charlson comorbidity score, frailty index, pregnancy, medical center area, and number of monovalent doses prior to index date.

[c] Medical center area dropped due to lack of model convergence.

[d] Charlson comorbidity score, frailty index, BMI, pregnancy, number of outpatient and virtual visits, number of emergency department visits, and number of monovalent doses prior to index date dropped due to lack of model convergence.

Supplementary Table 14. Vaccine effectiveness of mRNA-1273 bivalent vaccine vs. unvaccinated against infection and severe outcomes with XBB.1.5 subvariant

| | Test Positive | | Test Negative | | Odds Ratio (95% CI) | | VE (95% CI)[a] | |
|---|---|---|---|---|---|---|---|---|
| **Outcome** | **mRNA-1273 bivalent vaccinated (%)** | **Unvaccinated (%)** | **mRNA-1273 bivalent vaccinated (%)** | **Unvaccinated (%)** | **Unadjusted** | **Adjusted[b]** | **Unadjusted** | **Adjusted[b]** |
| SARS-CoV-2 infection[c] | 533 (44.9%) | 655 (55.1%) | 1485 (41.7%) | 2079 (58.3%) | 1.205 (1.028, 1.413) | 1.170 (0.988, 1.387) | -17.0% (-29.2%, -2.7%) | -14.6% (-27.9%, 1.2%) |
| ED/UC encounters[c] | 230 (39.2%) | 357 (60.8%) | 738 (41.9%) | 1023 (58.1%) | 0.847 (0.672, 1.066) | 0.837 (0.656, 1.069) | 15.3% (-6.2%, 32.8%) | 16.3% (-6.5%, 34.4%) |
| COVID-19 hospitalization[c,d] | 25 (38.5%) | 40 (61.5%) | 138 (70.8%) | 57 (29.2%) | 0.233 (0.124, 0.437) | 0.212 (0.100, 0.450) | 76.7% (56.3%, 87.6%) | 78.8% (55.0%, 90.0%) |
| COVID-19 hospital death | 1 (50.0%) | 1 (50.0%) | 4 (66.7%) | 2 (33.3%) | 0.577 (0.033, 10.25) | N/A | 42.3% (-90.2%, 96.7%) | N/A |

BMI= body mass index, ED/UC = emergency department/urgent care, OR = odds ratio, VE = vaccine effectiveness, N/A = not applicable
[a] Calculated as (1 – OR) x 100 when OR was ≤1, and ([1/OR] – 1) × 100 when OR was >1
[b] Applied conditional logistic models conditioned on matched pairs. Adjusted for time since history of SARS-CoV-2 infection, history of SARS-CoV-2 molecular test, number of outpatient and virtual visits, BMI, pregnancy, specimen type, and medical center area.
[c] Medical center area dropped due to lack of model convergence.
[d] Specimen type and pregnancy dropped due to lack of model convergence.

Supplementary Table 15. Vaccine effectiveness of mRNA-1273 bivalent vaccine vs. ≥2 monovalent mRNA vaccines against severe outcomes with SARS-CoV-2 variants among individuals without antiviral treatment

| | Test Positive | | Test Negative | | Odds Ratio (95% CI) | | rVE (95% CI)[a] | |
|---|---|---|---|---|---|---|---|---|
| Subvariant/Time since vaccination | mRNA-1273 bivalent vaccinated (%) | ≥2 doses monovalent mRNA vaccinated (%) | mRNA-1273 bivalent vaccinated (%) | ≥2 doses monovalent mRNA vaccinated (%) | Unadjusted | Adjusted [b,c] | Unadjusted | Adjusted [b,c] |
| BA.4/BA.5 | | | | | | | | |
| COVID-19 hospitalization | 23 (9.8%) | 212 (90.2%) | 184 (26.1%) | 521 (73.9%) | 0.296 (0.185, 0.473) | 0.359 (0.207, 0.621) | 70.4% (52.7%, 81.5%) | 64.1% (37.9%, 79.3%) |
| 14-60 days | 11 (4.9%) | 212 (95.1%) | 120 (18.7%) | 521 (81.3%) | 0.225 (0.119, 0.426) | 0.289 (0.146, 0.572) | 77.5% (57.4%, 88.1%) | 71.1% (42.8%, 85.4%) |
| 61-120 days | 11 (4.9%) | 212 (95.1%) | 62 (10.6%) | 521 (89.4%) | 0.436 (0.225, 0.844) | 0.544 (0.255, 1.159) | 56.4% (15.6%, 77.5%) | 45.6% (-13.7%, 74.5%) |
| 121-180 days | 0 (0.0%) | 212 (100.0%) | 2 (0.4%) | 521 (99.6%) | N/A | N/A | N/A | N/A |
| >180 days | 1 (0.5%) | 212 (99.5%) | 0 (0.0%) | 521 (100.0%) | N/A | N/A | N/A | N/A |
| COVID-19 hospital death [d,e] | 2 (12.5%) | 14 (87.5%) | 13 (27.1%) | 35 (72.9%) | 0.358 (0.069, 1.869) | 0.419 (0.031, 5.728) | 64.2% (-46.5%, 93.1%) | 58.1% (-82.5%, 96.9%) |
| XBB | | | | | | | | |
| COVID-19 hospitalization [d] | 37 (20.3%) | 145 (79.7%) | 173 (31.7%) | 373 (68.3%) | 0.562 (0.378, 0.837) | 0.504 (0.311, 0.817) | 43.8% (16.3%, 62.2%) | 52.9% (22.9%, 71.2%) |
| 14-60 days [d] | 2 (1.4%) | 145 (98.6%) | 24 (6.0%) | 373 (94.0%) | 0.214 (0.050, 0.919) | 0.142 (0.029, 0.691) | 78.6% (8.1%, 95.0%) | 85.8% (30.9%, 97.1%) |
| 61-120 days [d] | 6 (4.0%) | 145 (96.0%) | 46 (11.0%) | 373 (89.0%) | 0.336 (0.140, 0.803) | 0.205 (0.078, 0.538) | 66.4% (19.7%, 86.0%) | 79.5% (46.2%, 92.2%) |
| 121-180 days [d] | 12 (7.6%) | 145 (92.4%) | 57 (13.3%) | 373 (86.7%) | 0.542 (0.282, 1.039) | 0.495 (0.239, 1.026) | 45.8% (-3.7%, 71.8%) | 50.5% (-2.5%, 76.1%) |
| >180 days [d] | 17 (10.5%) | 145 (89.5%) | 46 (11.0%) | 373 (89.0%) | 0.951 (0.528, 1.712) | 0.978 (0.490, 1.952) | 4.9% (-41.6%, 47.2%) | 2.2% (-48.8%, 51.0%) |
| COVID-19 hospital death [d,e,f] | 4 (30.8%) | 9 (69.2%) | 16 (41.0%) | 23 (59.0%) | 0.649 (0.174, 2.413) | 0.695 (0.079, 6.142) | 35.1% (-58.6%, 82.6%) | 30.5% (-83.7%, 92.1%) |

BMI= body mass index, OR = odds ratio, rVE = relative vaccine effectiveness, N/A = not applicable
[a] Calculated as (1 – OR) x 100 when OR was ≤1, and ([1/OR] – 1) × 100 when OR was >1
[b] Applied conditional logistic models conditioned on matched pairs for hospitalization and hospital death outcomes. Models for time since vaccination analyses are unconditional logistic models, adjusted for age, sex, race/ethnicity, and month of specimen collection, in addition to the covariates listed below.
[c] Adjusted for days since last monovalent dose, time since history of SARS-CoV-2 infection, history of SARS-CoV-2 molecular test, number of outpatient and virtual visits, number of emergency department visits, BMI, Charlson comorbidity score, frailty index, pregnancy, and number of monovalent doses prior to index date.
[d] Pregnancy dropped due to lack of model convergence.
[e] BMI, Charlson comorbidity score, and frailty index dropped due to lack of model convergence.
[f] Number of monovalent doses prior to index date dropped due to lack of model convergence.

Supplementary Table 16. Vaccine effectiveness of mRNA-1273 bivalent vaccine vs. unvaccinated against severe outcomes with SARS-CoV-2 variants among individuals without antiviral treatment

| Subvariant/Time since vaccination | Test Positive | | Test Negative | | Odds Ratio (95% CI) | | VE (95% CI)[a] | |
|---|---|---|---|---|---|---|---|---|
| | mRNA-1273 bivalent vaccinated (%) | Unvaccinated (%) | mRNA-1273 bivalent vaccinated (%) | Unvaccinated (%) | Unadjusted | Adjusted [b,c] | Unadjusted | Adjusted [b,c] |
| BA.4/BA.5 | | | | | | | | |
| COVID-19 hospitalization | 23 (21.9%) | 82 (78.1%) | 149 (47.3%) | 166 (52.7%) | 0.245 (0.137, 0.438) | 0.231 (0.115, 0.461) | 75.5% (56.2%, 86.3%) | 76.9% (53.9%, 88.5%) |
| 14-60 days | 11 (11.8%) | 82 (88.2%) | 84 (33.6%) | 166 (66.4%) | 0.265 (0.134, 0.524) | 0.254 (0.119, 0.542) | 73.5% (47.6%, 86.6%) | 74.6% (45.8%, 88.1%) |
| 61-120 days | 11 (11.8%) | 82 (88.2%) | 60 (26.5%) | 166 (73.5%) | 0.371 (0.185, 0.744) | 0.376 (0.175, 0.808) | 62.9% (25.6%, 81.5%) | 62.4% (19.2%, 82.5%) |
| 121-180 days | 0 (0.0%) | 82 (100.0%) | 5 (2.9%) | 166 (97.1%) | N/A | N/A | N/A | N/A |
| >180 days | 1 (1.2%) | 82 (98.8%) | 0 (0.0%) | 166 (100.0%) | N/A | N/A | N/A | N/A |
| COVID-19 hospital death [d,e] | 2 (22.2%) | 7 (77.8%) | 10 (37.0%) | 17 (63.0%) | 0.518 (0.097, 2.755) | 0.541 (0.099, 2.947) | 48.2% (-63.7%, 90.3%) | 45.9% (-66.1%, 90.1%) |
| XBB | | | | | | | | |
| COVID-19 hospitalization [d] | 37 (44.6%) | 46 (55.4%) | 175 (70.3%) | 74 (29.7%) | 0.309 (0.179, 0.532) | 0.290 (0.156, 0.538) | 69.1% (46.8%, 82.1%) | 71.0% (46.2%, 84.4%) |
| 14-60 days [d] | 2 (4.2%) | 46 (95.8%) | 30 (28.8%) | 74 (71.2%) | 0.107 (0.024, 0.470) | 0.073 (0.015, 0.355) | 89.3% (53.0%, 97.6%) | 92.7% (64.5%, 98.5%) |
| 61-120 days [d] | 6 (11.5%) | 46 (88.5%) | 55 (42.6%) | 74 (57.4%) | 0.175 (0.070, 0.440) | 0.142 (0.052, 0.389) | 82.5% (56.0%, 93.0%) | 85.8% (61.1%, 94.8%) |
| 121-180 days [d] | 12 (20.7%) | 46 (79.3%) | 46 (38.3%) | 74 (61.7%) | 0.420 (0.201, 0.874) | 0.378 (0.163, 0.877) | 58.0% (12.6%, 79.9%) | 62.2% (12.3%, 83.7%) |
| >180 days [d] | 17 (27.0%) | 46 (73.0%) | 44 (37.3%) | 74 (62.7%) | 0.622 (0.318, 1.214) | 0.581 (0.263, 1.282) | 37.8% (-17.7%, 68.2%) | 41.9% (-22.0%, 73.7%) |
| COVID-19 hospital death [d,e] | 4 (80.0%) | 1 (20.0%) | 13 (86.7%) | 2 (13.3%) | 0.577 (0.033, 10.25) | 0.690 (0.035, 13.55) | 42.3% (-90.2%, 96.7%) | 31.0% (-92.6%, 96.5%) |

BMI= body mass index, OR = odds ratio, VE = vaccine effectiveness, N/A = not applicable
[a] Calculated as (1 – OR) x 100 when OR was ≤1, and ([1/OR] – 1) × 100 when OR was >1
[b] Applied conditional logistic models conditioned on matched pairs for hospitalization and hospital death outcomes. Models for time since vaccination analyses are unconditional logistic models, adjusted for age, sex, race/ethnicity, and month of specimen collection, in addition to the covariates listed below.
[c] Adjusted for time since history of SARS-CoV-2 infection, history of SARS-CoV-2 molecular test, number of outpatient and virtual visits, specimen type and BMI.
[d] Specimen type dropped due to lack of model convergence.
[e] BMI and time since history of SARS-CoV-2 infection dropped due to lack of model convergence.

Supplementary Figure 1. Flow chart for Moderna bivalent vaccine test-negative design study population

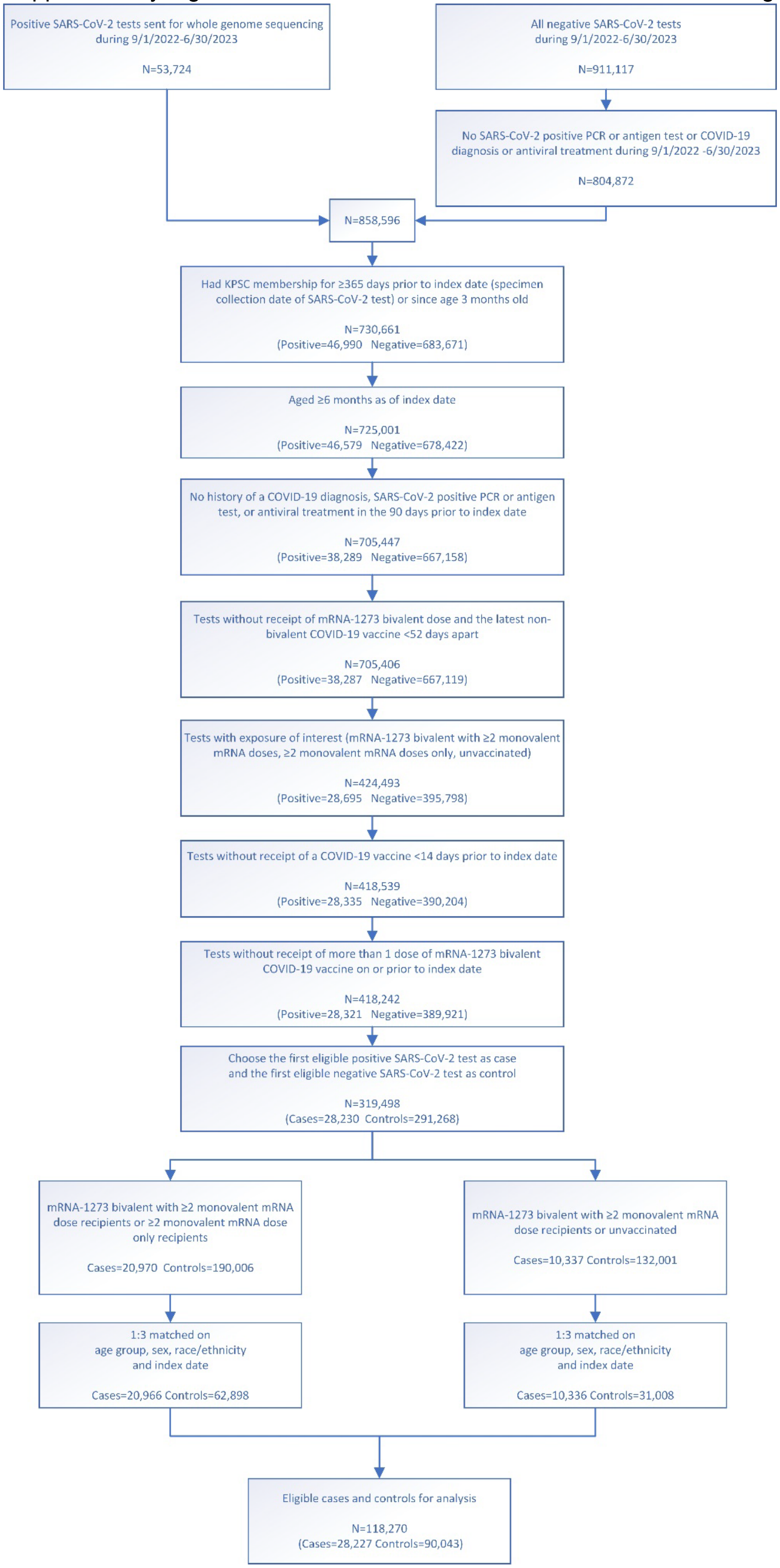

Supplementary Figure 2. Distribution of SARS-CoV-2 variants by vaccination status

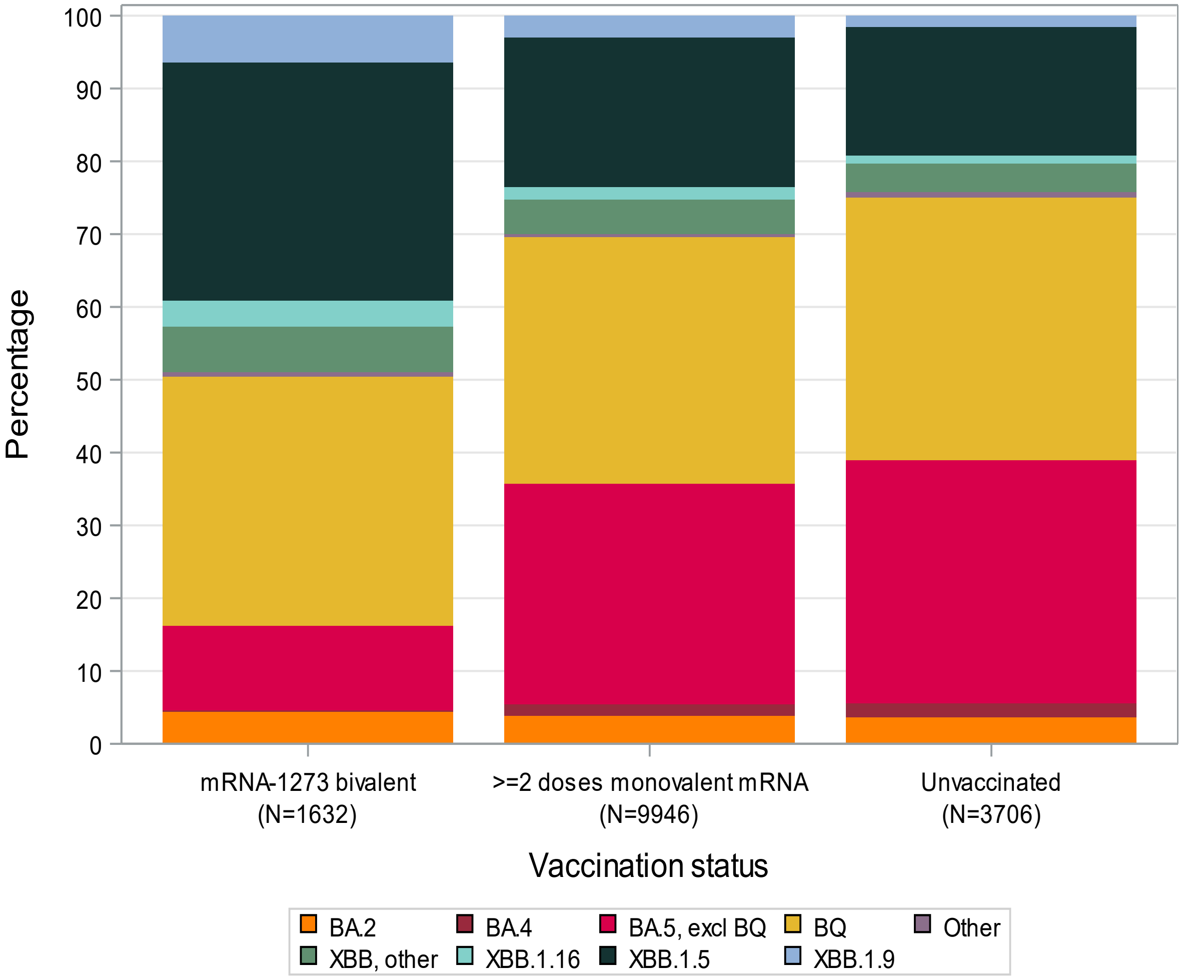

Supplementary Figure 3. Distribution of SARS-CoV-2 variants by month of specimen collection

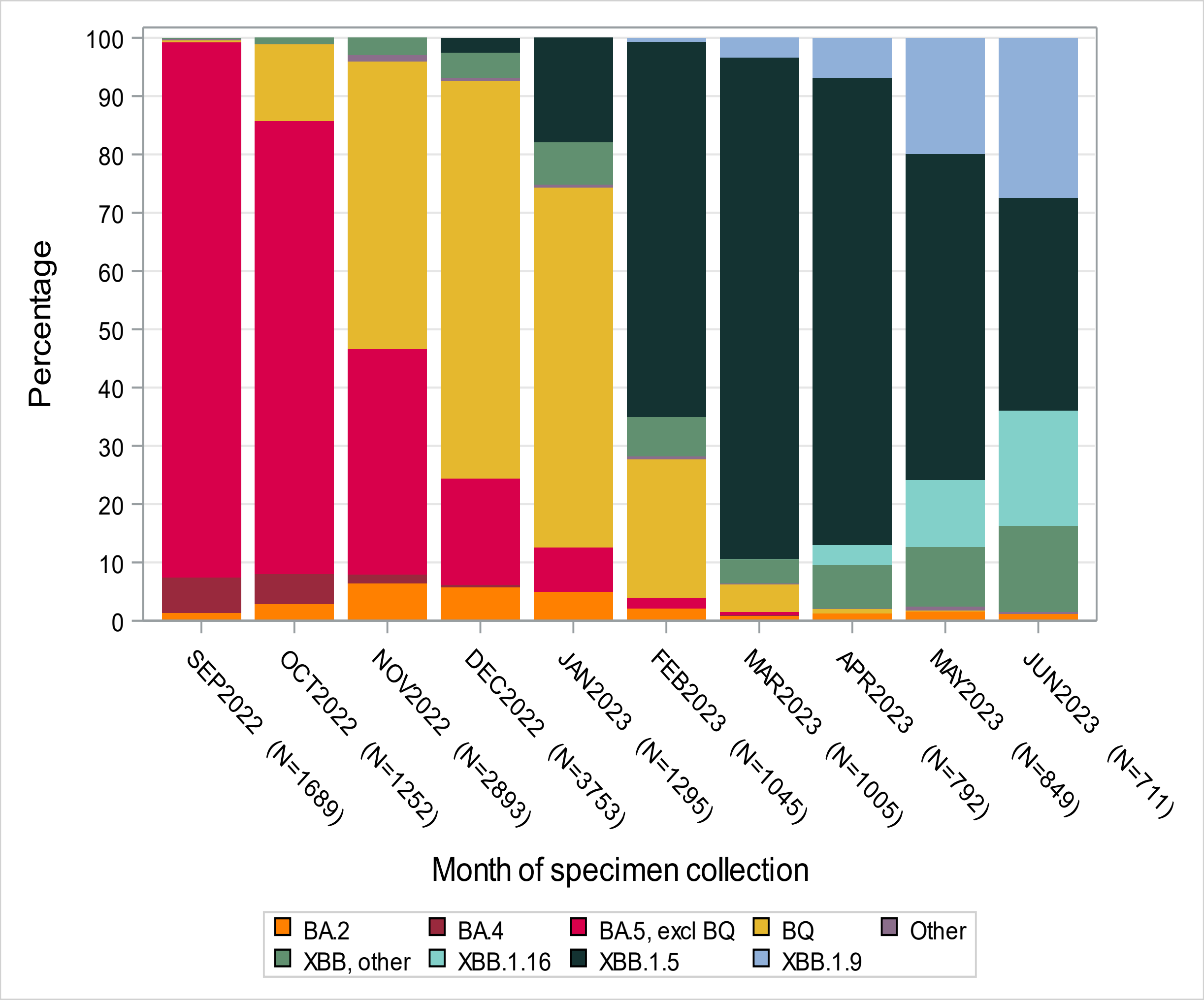
